# Comparative safety and effectiveness of Pfizer BA.4-5 versus Sanofi during the spring 2023 COVID-19 booster vaccination programme in England: a matched cohort study in OpenSAFELY-TPP

**DOI:** 10.1101/2024.03.15.24304277

**Authors:** Colm D Andrews, Em Prestige, Edward PK Parker, Venexia Walker, Tom Palmer, Andrea L Schaffer, Amelia CA Green, Helen J Curtis, Alex Walker, Rebecca M Smith, Christopher Wood, Chris Bates, Amir Mehrkar, Brian MacKenna, Sebastian CJ Bacon, Ben Goldacre, Miguel A Hernán, Jonathan AC Sterne, the OpenSAFELY collaborative, William J Hulme

## Abstract

**Introduction:** The spring 2023 COVID-19 booster vaccination programme in England used both Pfizer BA.4-5 and Sanofi vaccines. All people aged 75 years or over and the clinically vulnerable were eligible to receive a booster dose. Direct comparisons of the effectiveness of these two vaccines in boosting protection against severe COVID-19 events have not been made in trials or observational data.

**Methods:** With the approval of NHS England, we used the OpenSAFELY-TPP database to compare effectiveness of the Pfizer BA.4-5 and Sanofi vaccines during the spring 2023 booster programme, between 1 April and 30 June 2023. We investigated two cohorts separately: those aged 75 or over (75+); and those aged 50 or over and clinically vulnerable (CV). In each cohort, vaccine recipients were matched on date of vaccination, COVID-19 vaccine history, age, and other characteristics. Effectiveness outcomes were COVID-19 hospital admission, COVID-19 critical care admission, and COVID-19 death up to 16 weeks after vaccination. Safety outcomes were pericarditis and myocarditis up to 4 weeks after vaccination. We report the cumulative incidence of each outcome, and compare safety and effectiveness using risk differences (RD), relative risks (RR), and incidence rate ratios (IRRs).

**Results:** 492,642 people were 1-1 matched in the CV cohort, and 673,926 in the 75+ cohort, contributing a total of 7,423,251 and 10,173,230 person-weeks of follow-up, respectively. The incidence of COVID-19 hospital admission was higher for Sanofi than for Pfizer BA.4-5. In the CV cohort, 16-week risks per 10,000 people were 22.3 (95%CI 20.4 to 24.3) for Pfizer BA.4-5 and 26.4 (24.4 to 28.7) for Sanofi, with an IRR of 1.19 (95%CI 1.06 to 1.34). In the 75+ cohort, these were 17.5 (16.1 to 19.1) for Pfizer BA.4-5 and 20.4 (18.9 to 22.1) for Sanofi, with an IRR of 1.18 (1.05-1.32). These findings were similar across all pre-specified subgroups. More severe COVID-19 related outcomes (critical care admission and death), and safety outcomes at 4 weeks, were rare in both vaccines so we could not reliably compare effectiveness of the two vaccines.

**Conclusion:** This observational study comparing effectiveness of Pfizer BA.4-5 and Sanofi vaccine during the spring 2023 programme in England in the two main eligible cohorts – people aged 75 and over and in clinically vulnerable people – found some evidence of superior effectiveness against COVID-19 hospital admission for Pfizer BA.4-5 compared with Sanofi within 16 weeks after vaccination.

## Background

The spring 2023 COVID-19 booster vaccination programme in England offered COVID-19 vaccination to those aged 75 years or over, resident in a care home, and those who are immunosuppressed, based on guidance from the Joint Committee for Vaccination and Immunisation (JCVI).(1–3) Both the Pfizer BA.4-5 vaccine (authorised November 2022 (4)) and the Sanofi vaccine (authorised December 2022 (5)) were used during this programme. This allows direct comparisons of their safety and effectiveness against severe COVID-19 disease – something not investigated in randomised trials.

We used the OpenSAFELY-TPP database, covering around 45% of English primary care practices and linked to hospital and death records, to compare safety and effectiveness of the Pfizer BA.4-5 and Sanofi vaccines received during the spring 2023 booster programme.

## Methods

### Data source

All data were linked, stored and analysed securely using the OpenSAFELY platform: https://opensafely.org/, as part of the NHS England OpenSAFELY COVID-19 service. Data include pseudonymised data such as coded diagnoses, medications and physiological parameters. No free text data are included. All code is shared openly for review and re-use under MIT open license (https://github.com/opensafely/comparative-booster-spring2023). Detailed pseudonymised patient data is potentially re-identifiable and therefore not shared. Primary care records managed by the GP software provider, TPP were linked, using NHS numbers, to emergency department attendance and in-patient hospital spell records via NHS Digital’s Hospital Episode Statistics (HES), national coronavirus testing records via the Second Generation Surveillance System (SGSS), and national death registry records from the Office for National Statistics (ONS). COVID-19 vaccination history and health and social care worker status is available in the GP record directly via the National Immunisation Management System (NIMS).

### Eligibility criteria

We considered two cohorts: all adults aged 75 years or over (“75+”), and all adults aged 50 or over who were considered clinically vulnerable (“CV”). These groups are not mutually exclusive. We only considered those aged 50 years or over because the Sanofi vaccine was rarely offered to younger people. Clinical vulnerability encompasses various chronic or immunosuppressive conditions and medications as set out in the Green Book chapter 14a (6), and defined in the supplementary materials.

In each group, any person receiving a Pfizer BA.4-5 or Sanofi vaccine dose between 1 April 2023 and 30 June 2023 inclusive was considered. People were eligible if they: were registered at a GP practice using TPP’s SystmOne clinical information system at the time of vaccination; had received at least two prior COVID-19 vaccine doses; were not a health or social care worker and not receiving end-of-life care; had no evidence of SARS-CoV-2 infection or COVID-19 disease during the 28 days prior to vaccination; were not a hospital in-patient at the time of vaccination; and had complete information on sex, deprivation, and Sustainability and Transformation Partnership (STP, a geographical grouping of NHS and Local Authorities).

### Matching

Pfizer BA.4-5 and Sanofi vaccine recipients were matched 1-1 without replacement, on the following characteristics: date of vaccination (3-day caliper); number of previous COVID-19 vaccine doses (grouped as 2 to 4, 5, or 6 or more); time since previous COVID-19 vaccine dose (28-day caliper); sex (male or female); age (3-year caliper and within age groups 50-64, 65-75, 75-79, 80-84, 85+); Index of Multiple Deprivation (IMD, within a 5000 IMD-rank caliper); STP as a surrogate for geographical region; any evidence of prior SARS-CoV-2 infection (including any of: positive SARS-CoV-2 test, infection as documented in primary care, COVID-19 hospital attendance or admission); and morbidity count (grouped as 0, 1, or 2 or more conditions from the following list: diabetes, BMI over 40kg/m^2^, chronic heart disease, chronic kidney disease, chronic liver disease, chronic respiratory disease or severe asthma, chronic neurological disease, cancer within 3 years). Clinical vulnerability was an additional matching factor in the 75+ cohort.

Age was calculated as of 30 June 2023, to allow for a degree of operational flexibility in who was offered vaccination during the programme. All other characteristics were calculated as of the day before vaccination. In the analysis cohorts there are no missing values for any characteristics as people with missing values for sex, IMD or STP were excluded, and remaining characteristics were defined by the presence or absence of clinical codes or events in the health record. The supplementary materials provide more information on how these characteristics were defined.

### Outcomes and follow-up

We considered three effectiveness outcomes. COVID-19 hospital admission was identified using Secondary Use Service (SUS) in-patient hospital spell records with International Statistical Classification of Diseases and Related Health Problems 10th Revision (ICD-10) codes U07.1, U07.2, or U.109 as the primary or non-primary reason for admission. COVID-19 critical care admissions were any COVID-19 hospital spell with at least one day spent in critical care (not limited to the first hospitalisation spell after vaccination). COVID-19 death was defined as COVID-19 ICD-10 codes (as before) mentioned anywhere on the death certificate (i.e., as an underlying or contributing cause of death). We also report on non-COVID-19 deaths.

We considered two safety outcomes. Pericarditis within 28 days of vaccination was identified using SUS emergency department records with pericarditis listed as the discharge diagnosis (SNOMED), and using ICD-10 coded hospital admission and death records. Myocarditis was identified similarly. Details of the codelists used to define these outcomes are available in the supplementary materials.

Each person was followed from receipt of vaccination (time zero) until the outcome of interest, with censoring at death, practice de-registration, or 31 August 2023. Outcomes were observed for up to 16 weeks for effectiveness outcomes, and up to 4 weeks for safety outcomes.

### Statistical Analysis

Baseline characteristics of each vaccine group were tabulated, with between-group balance examined using standardised mean differences. We estimated the cumulative incidence of each outcome in each vaccine group using the Kaplan-Meier (KM) estimator. We estimated cumulative risk differences (RDs) and risk ratios (RRs) comparing the vaccine groups for each outcome. We also estimated Incidence Rate Ratios (IRRs) for Sanofi versus Pfizer BA.4-5 recipients over the full follow-up duration, and within period-specific intervals defined by splits on weeks 1, 2, 4, 8, 12, and 16.

95% confidence limits for the cumulative incidences were derived using standard errors on the log-KM scale. Confidence limits for the risk differences were derived from the sum of squares of the standard errors on the KM scale, using Greenwood’s formula. Confidence limits for the risk ratios and incidence rate ratios were derived from the sum of squares of the standard errors on the log-KM scale.

### Subgroup and secondary analyses

We estimated comparative effectiveness separately in the following subgroups: age band in the CV group only (50-64, 65-75, 75-79, 80-84, 85+); clinical vulnerability in the 75+ group only; prior COVID-19 vaccine count. We used χ^2^ tests to examine evidence for heterogeneity in estimates between subgroups.

### Disclosure control

To satisfy strict re-identification minimisation requirements for statistical outputs from OpenSAFELY’s Trusted Research Environment, counts were rounded to the nearest 3, 9, 15, and so on. Plots of cumulative event counts and the Kaplan-Meier cumulative incidence estimates were rounded such that each increment is based on at least 6 events. Event rates, risk differences, and risk ratios were derived from these rounded estimates.

### Software, code, and reproducibility

Data management and analyses were conducted using OpenSAFELY tools, Python version 3.8.10 and R version 4.0.2. Code for data management and analysis, as well as codelists, are archived online (https://github.com/opensafely/comparative-booster-spring2023). Codelists are available at https://www.opencodelists.org/. The supplementary materials provide further details of codelists and data sources used for all variables in the study. Detailed pseudonymised patient data is potentially re-identifiable and therefore not shared.

## Results

### Study population and matching

We identified 2,023,962 people aged 50 years or over who were recorded as having received a COVID-19 vaccine between 1 April 2023 and 30 June 2023 whilst registered at a TPP practice. This included 976,422 (48.2%) Pfizer BA.4-5 and 963,294 (47.6%) Sanofi recipients.

We identified 1,510,158 people in the CV cohort, and of these 736,098 (48.7%) received Pfizer BA.4-5 and 710,526 (47.0%) received Sanofi, with 679,314 (92.3%) and 647,244 (91.1%) eligible for matching respectively (Figure 1a).

**Figure 1a:**
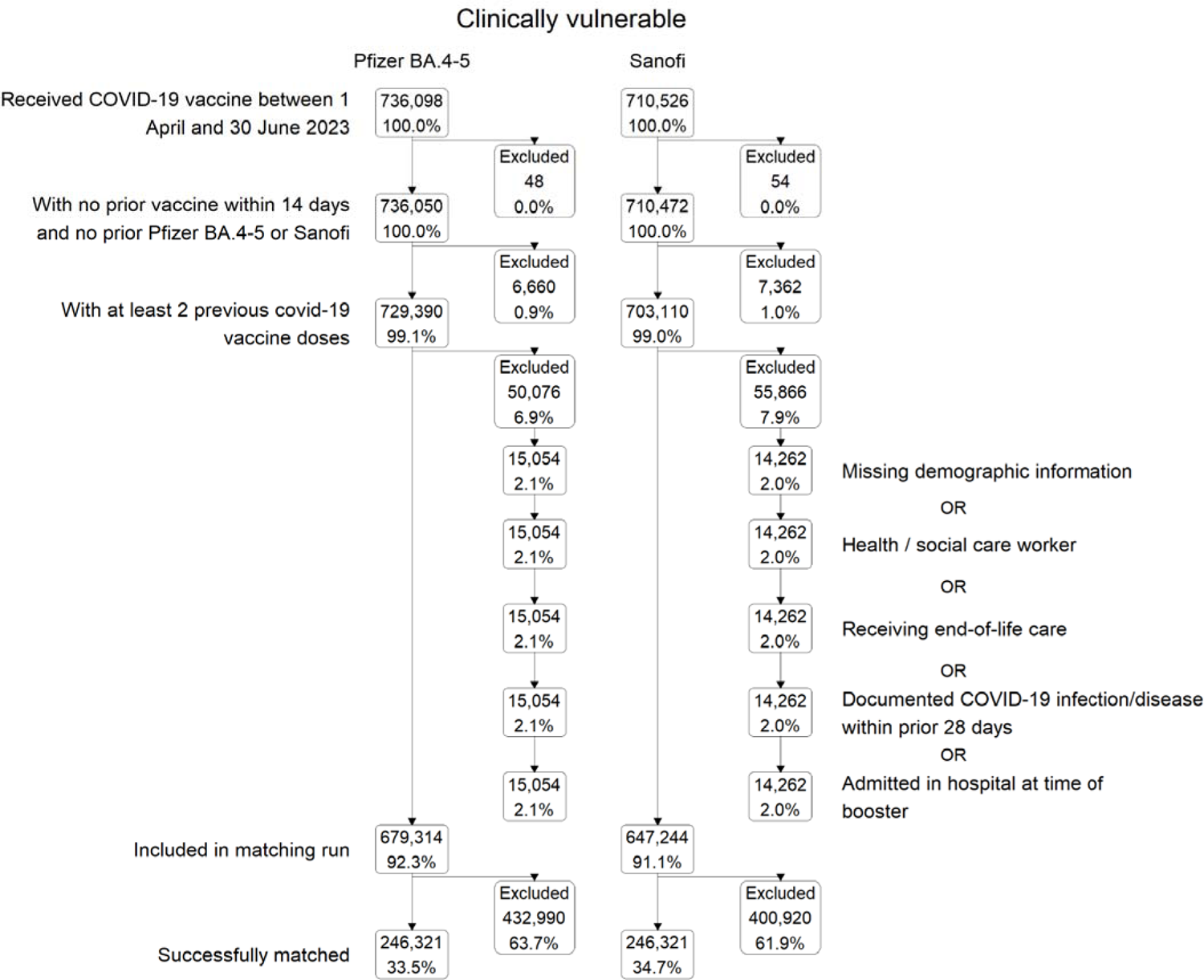
Flow of participants into the CV cohort

1,782,636 people were in the 75+ cohort, and of these 770,610 (43.2%) received Pfizer BA.4-5 and 946,038 (53.1%) received Sanofi, with 724,092 (94.0%) and 876,174 (92.6%) eligible for matching respectively (Figure 1b).

**Figure 1b:**
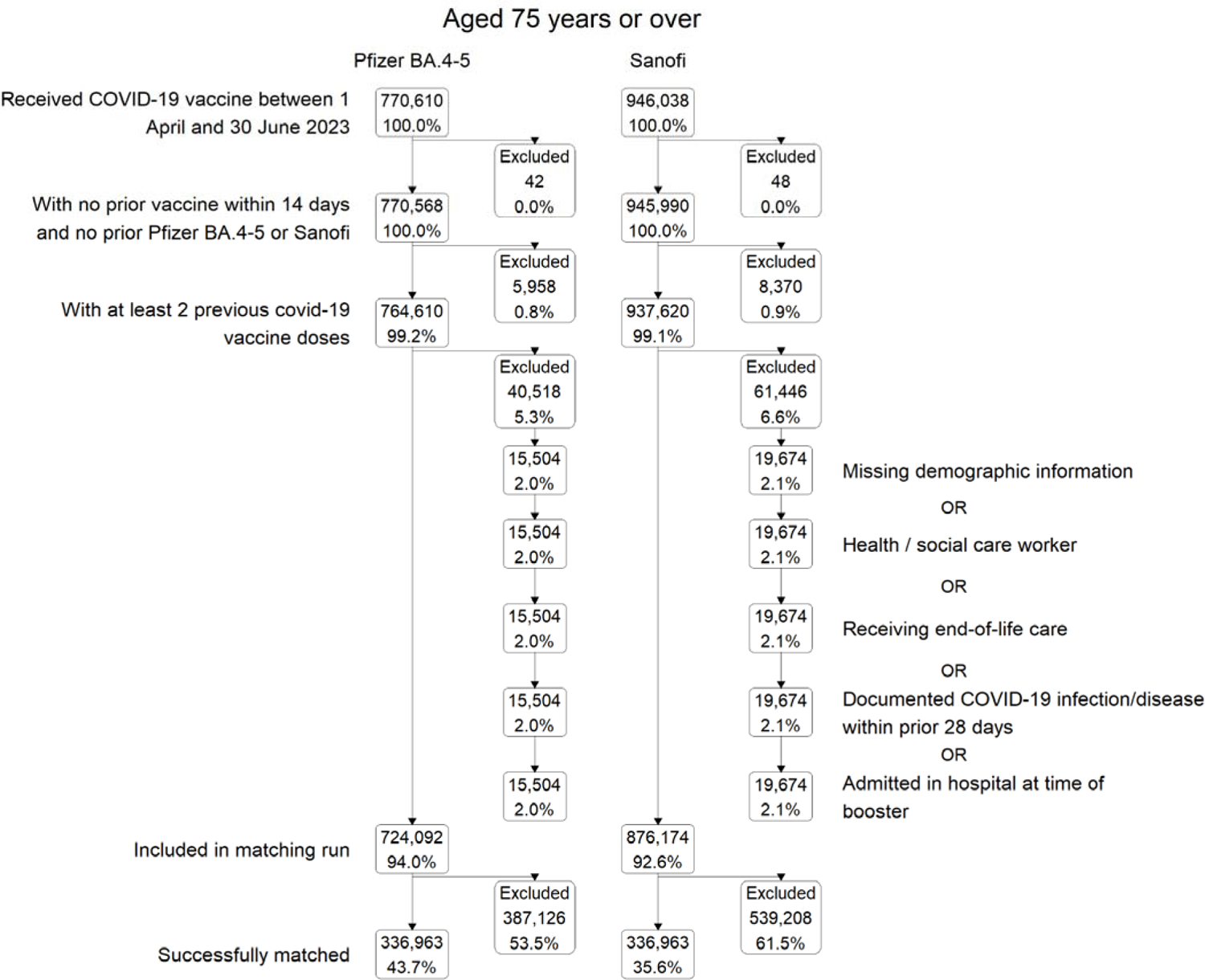
Flow of participants into the 75+ cohort

**Figure 2a:**
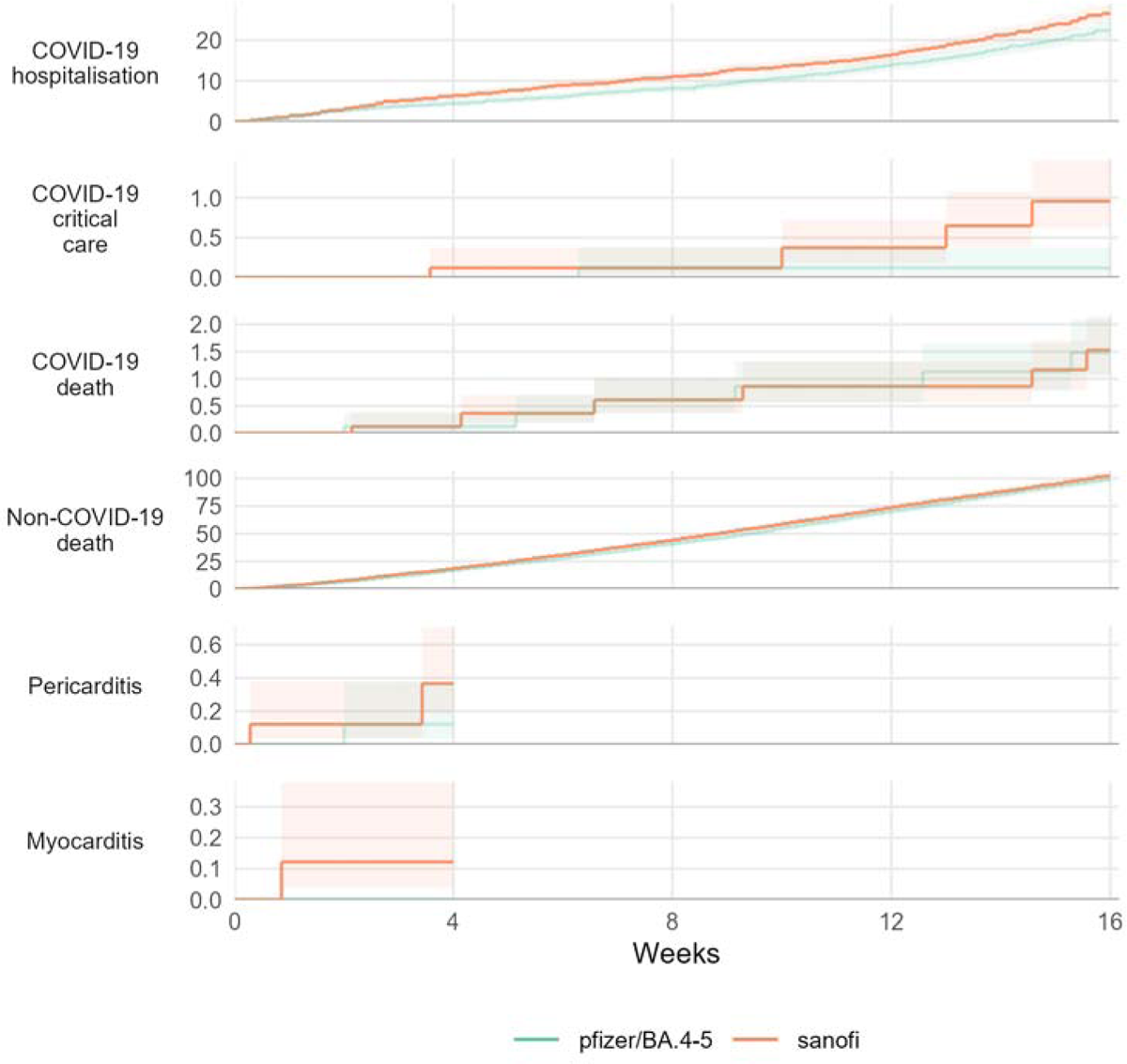
Estimates in the CV cohort of cumulative risk per 10,000.

**Figure 2b:**
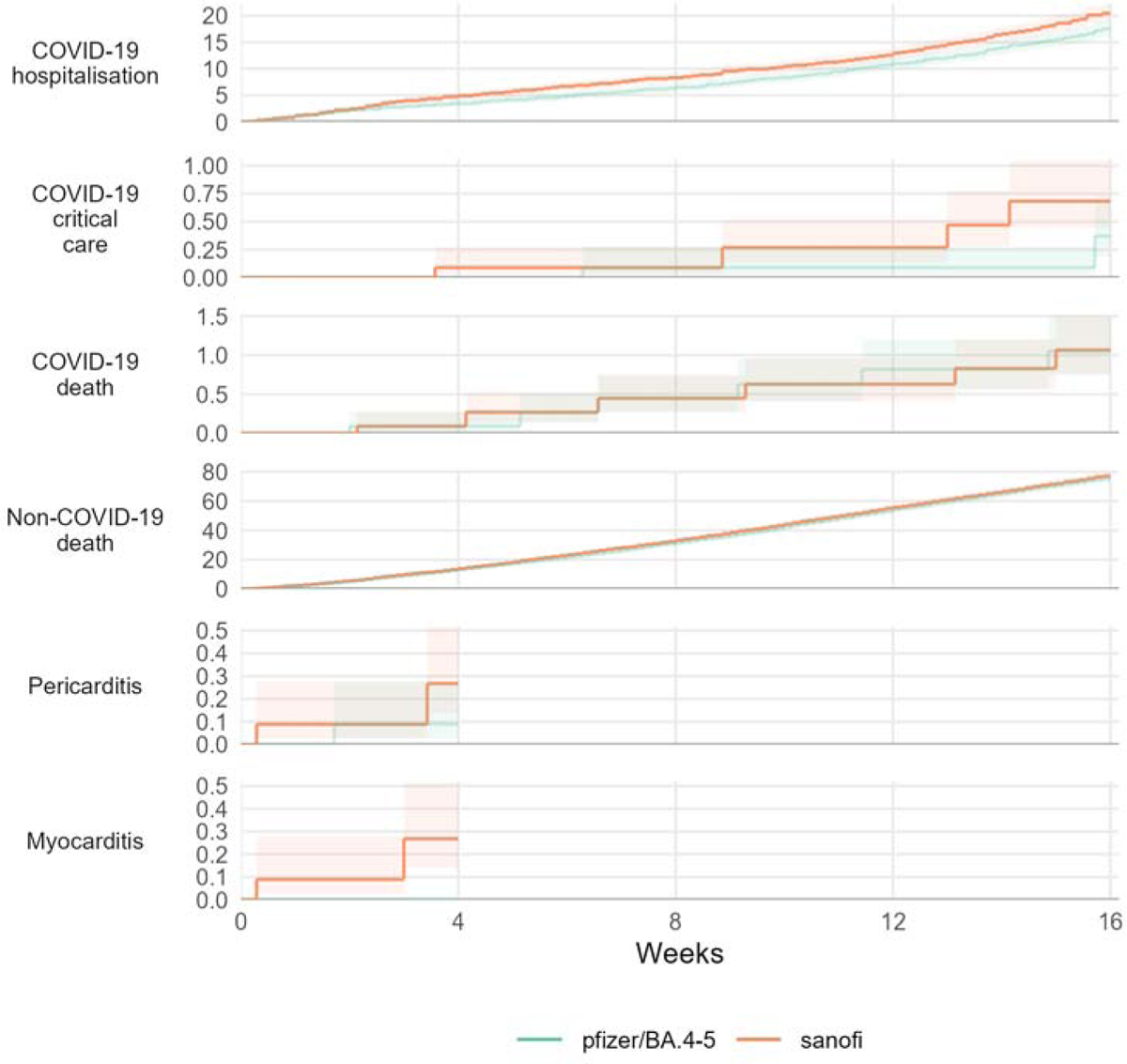
Estimates in the 75+ cohort of cumulative risk per 10,000.

Sanofi was more likely to be administered in April 2023, and Pfizer BA.4-5 more likely in May and June 2023 (Figure S1a and S1b). Prior to matching, Sanofi recipients were on average older, more likely to be recorded as resident in a care or nursing home, and less likely to have received fewer than 5 prior COVID-19 vaccine doses compared with Pfizer BA.4-5 recipients in both cohorts. Sanofi recipients were less likely than Pfizer BA.4-5 recipients to have previously received the Pfizer/BA.1 vaccine during an earlier booster programme and more likely to have received Moderna/Omicron. Sex, deprivation levels, and ethnicity were similar in both groups prior to matching (supplementary Tables S1a and S1b).

In the CV cohort, 246,321 people in each vaccine group (total 492,642) were matched, representing 36.3% and 38.1% of eligible Pfizer and Sanofi recipients respectively (Figure 1a, Figure S1a). In the 75+ cohort, 336,963 were matched in each vaccine group (total 673,926), representing 46.5% and 38.5% of eligible Pfizer and Sanofi recipients respectively (Figure 1b, Figure S1b).

After matching, characteristics at the start of follow-up were well-balanced between groups (Table 1) with standardised mean differences consistently under 0.05 (supplementary Figures S2a and S2b), including for those characteristics not used for matching.

**Table 1:**
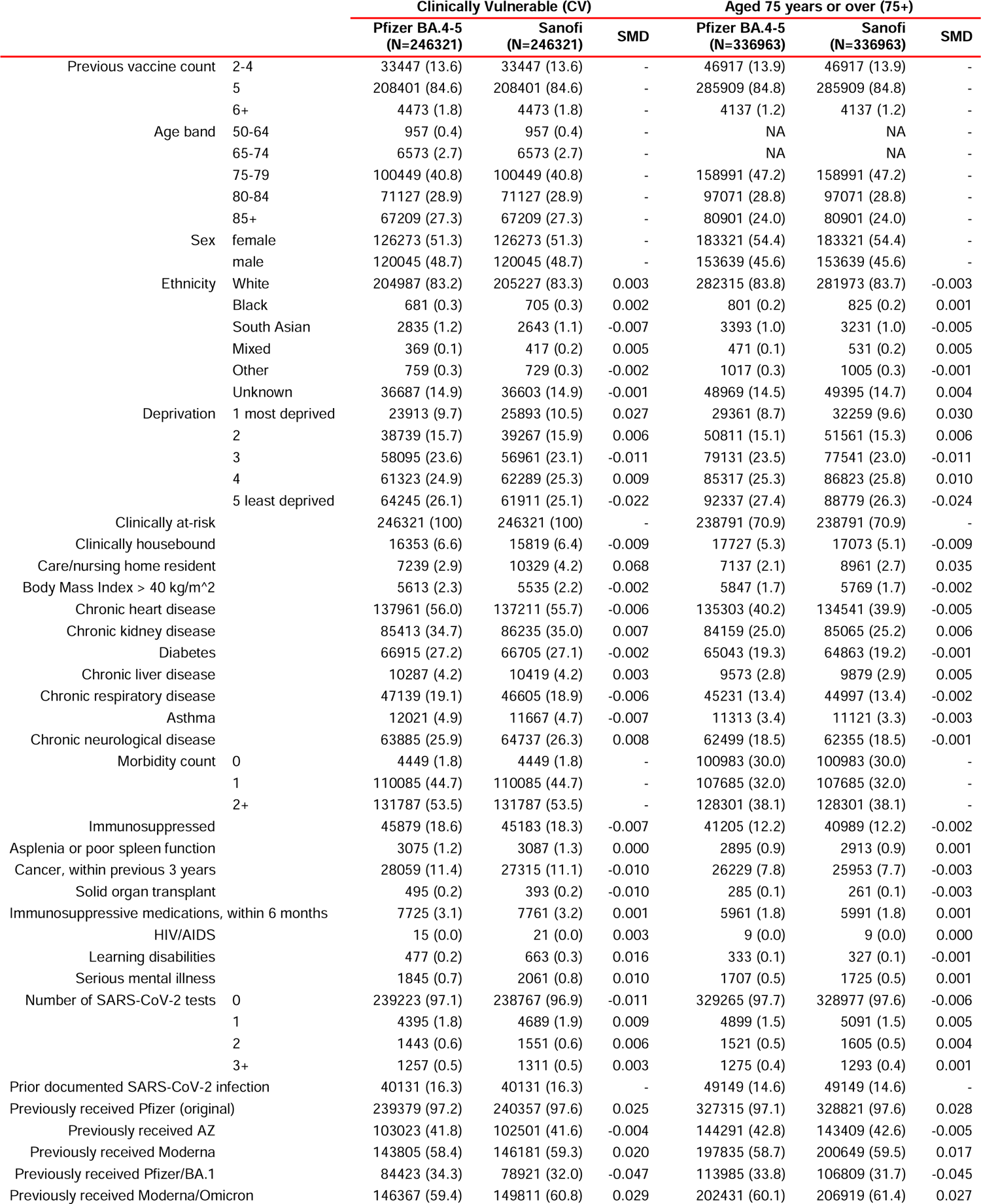
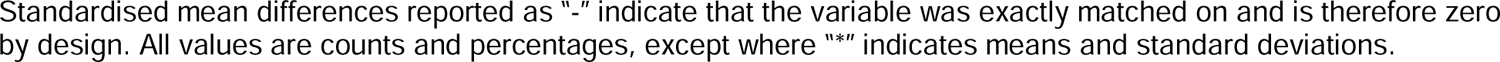
Baseline characteristics before and after matching.

### Estimated comparative effectiveness

#### Clinically Vulnerable cohort

At 16 weeks there were 1,098 COVID-19 hospital admissions, 24 COVID-19 critical care admissions, and 66 COVID-19 deaths (Table 2). The 16-week risks (cumulative incidence) per 10,000 people of COVID-19 hospital admissions were 22.3 (95%CI 20.4 to 24.3) for those receiving Pfizer BA.4-5 and 26.4 (24.4 to 28.7) for Sanofi (risk difference 4.15 (95%CI 1.25 to 7.05) where values greater than zero favour Pfizer) (Table 2; Figure 3). The corresponding 16-week risks per 10,000 people for COVID-19 critical care admission were 0.12 (0.040 to 0.38) for Pfizer BA.4-5 and 0.96 (0.62 to 1.47) for Sanofi (risk difference 0.83 (0.40 to 1.27)). The 16-week risks per 10,000 people of COVID-19 death were 1.48 (1.05 to 2.09) for Pfizer BA.4-5 and 1.53 (1.08 to 2.16) for Sanofi (risk difference 0.044 (−0.69 to 0.78)). For non-COVID-19 death, these were 98.8 (94.8 to 102.9) for Pfizer BA.4-5 and 102.1 (98.0 to 106.3) for Sanofi (risk difference 3.28 (−2.50 to 9.07)).

**Figure 3:**
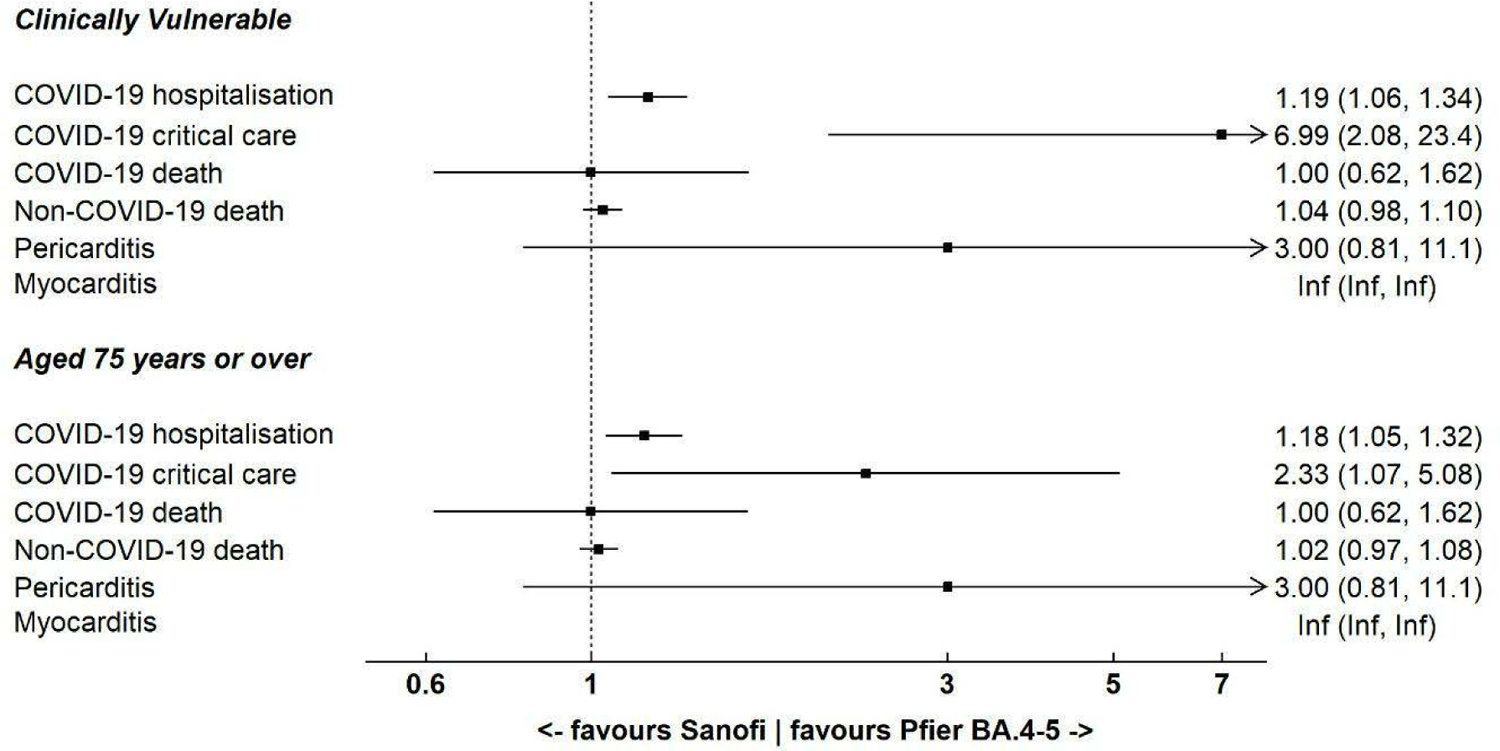
Estimated incidence rate ratios at comparing the safety (4-weeks) and effectiveness (16-weeks) of Sanofi and Pfizer BA.4-5, overall and within subgroups,.

**Table 2:** Estimates at 16-week follow-up of risk per 10,000 people in each vaccine group, and corresponding risk differences, overall and within subgroups Counts and survival estimates are based on values rounded up to the nearest 6*n* − 3 for disclosure control.

| Outcome | Pfizer BA.4-5 |  | Sanofi |  | Comparison (reference = Pfizer BA.4-5) |  |  |
| --- | --- | --- | --- | --- | --- | --- | --- |
|  | Events / Person-weeks | Risk per 10,000 people (95% CI) | Events / Person-weeks | Risk per 10,000 people (95% CI) | Risk difference per 10,000 people (95% CI) | Risk ratio (95% CI) | Incidence rate ratio (95% CI) |
| **Clinically Vulnerable** |  |  |  |  |  |  |  |
| COVID-19 hospitalisation | 501 / 3,705,754 | 22.3 (20.4 to 24.3) | 597 / 3,711,210 | 26.4 (24.4 to 28.7) | 04.2 (1.3 to 7.1) | 1.19 (1.05-1.34) | 1.19 (1.06-1.34) |
| COVID-19 critical care | 3 / 3,708,523 | 0.12 (0.04 to 0.38) | 21 / 3,714,624 | 0.96 (0.62 to 1.5) | 0.83 (0.40 to 1.3) | 7.80 (2.32-26.16) | 6.99 (2.08-23.43) |
| COVID-19 death | 33 / 3,708,546 | 1.5 (1.1 to 2.1) | 33 / 3,714,705 | 1.5 (1.1 to 2.2) | 0.04 (-0.69 to 0.78) | 1.03 (0.63-1.68) | 1.00 (0.62-1.62) |
| Non-COVID-19 death | 2,277 / 3,708,546 | 98.8 (94.8 to 103) | 2,361 / 3,714,705 | 102 (98.0 to 106) | 3.3 (-2.5 to 9.1) | 1.03 (0.98-1.09) | 1.04 (0.98-1.10) |
| Pericarditis | 3 / 983,527 | 0.12 (0.04 to 0.38) | 9 / 983,193 | 0.37 (0.19 to 0.70) | 0.24 (-0.03 to 0.52) | 3.00 (0.81-11.09) | 3.00 (0.81-11.09) |
| Myocarditis | 0 / 983,531 | 0.00 (-) | 3 / 983,195 | 0.12 (0.04 to 0.38) | 0.12 (-0.02 to 0.26) | Inf (-) | Inf (-) |
| **Aged 75 years or over** |  |  |  |  |  |  |  |
| COVID-19 hospitalisation | 537 / 5,078,234 | 17.5 (16.1 to 19.1) | 633 / 5,088,333 | 20.4 (18.9 to 22.1) | 2.9 (0.73 to 5.1) | 1.17 (1.04-1.31) | 1.18 (1.05-1.32) |
| COVID-19 critical care | 9 / 5,081,180 | 0.37 (0.19 to 0.72) | 21 / 5,091,929 | 0.68 (0.44 to 1.0) | 0.31 (-0.07 to 0.69) | 1.83 (0.83-4.05) | 2.33 (1.07-5.08) |
| COVID-19 death | 33 / 5,081,214 | 1.1 (0.75 to 1.5) | 33 / 5,092,016 | 1.1 (0.76 to 1.5) | 0.01 (-0.50 to 0.53) | 1.01 (0.62-1.65) | 1.00 (0.62-1.62) |
| Non-COVID-19 death | 2,379 / 5,081,214 | 75.4 (72.4 to 78.5) | 2,439 / 5,092,016 | 77.1 (74.1 to 80.2) | 1.7 (-2.6 to 6.0) | 1.02 (0.97-1.08) | 1.02 (0.97-1.08) |
| Pericarditis | 3 / 1,345,724 | 0.09 (0.03 to 0.28) | 9 / 1,345,418 | 0.27 (0.14 to 0.51) | 0.18 (-0.02 to 0.38) | 3.00 (0.81-11.09) | 3.00 (0.81-11.08) |
| Myocarditis | 0 / 1,345,728 | 0.00 (-) | 9 / 1,345,421 | 0.27 (0.14 to 0.51) | 0.27 (0.09 to 0.44) | Inf (-) | Inf (-) |

IRRs (where values greater than one favour Pfizer) in the 16-week period after vaccination were 1.19 (95%CI 1.06 to 1.34) for COVID-19 hospital admissions, 6.99 (2.08-23.43) for COVID-19 critical care admission, 1.00 (0.62-1.62) for COVID-19 death, and 1.04 (0.98-1.10) for non-COVID-19 death (Figure 3).

### 75+ cohort

By 16 weeks there were 1,170 COVID-19 hospital admissions, 30 COVID-19 critical care admissions, and 66 COVID-19 deaths (Table 2). The 16-week risks (cumulative incidence) per 10,000 people of COVID-19 hospital admissions were 17.5 (95%CI 16.1 to 19.1) for those receiving Pfizer BA.4-5 and 20.4 (18.9 to 22.1) for Sanofi (risk difference 2.92 (95%CI 0.73 to 5.11)) (Table 2; Figure 3). The corresponding 16-week risks per 10,000 people for COVID-19 critical care admission were 0.37 (0.19 to 0.72) for Pfizer BA.4-5 and 0.68 (0.44 to 1.05) for Sanofi (risk difference 0.31 (−0.074 to 0.69)). The 16-week risks per 10,000 people of COVID-19 death were 1.05 (0.75 to 1.48) for Pfizer BA.4-5 and 1.07 (0.76 to 1.50) for Sanofi (risk difference 0.013 (−0.50 to 0.53)). For non-COVID-19 death, these were 75.4 (72.4 to 78.5) for Pfizer BA.4-5 and 77.1 (74.1 to 80.2) for Sanofi (risk difference 1.71 (−2.60 to 6.02)).

IRRs in the 16-week period after vaccination were 1.18 (95%CI 1.05 to 1.32) for COVID-19 hospital admission, 2.33 (1.07-5.08) for COVID-19 critical care admission, 1.00 (0.62-1.62) for COVID-19 death, and 1.02 (0.97-1.08) for non-COVID-19 death (Figure 3).

Period-specific IRRs show that, in both cohorts during the first week after vaccination, event rates were similar between vaccine groups across all effectiveness outcomes (Supplementary Figure S4).

### Estimated comparative safety

In the CV cohort, there were 3 and 9 pericarditis events within 4 weeks after vaccination for Pfizer BA.4-5 and Sanofi recipients, respectively. This corresponded to a cumulative incidence per 10,000 people of 0.12 (95%CI 0.039 to 0.38) for Pfizer BA.4-5 and 0.37 (0.19 to 0.70) for Sanofi (risk difference 0.24 (95%CI −0.032 to 0.52)). For myocarditis, there were 0 events for Pfizer BA.4-5 and 3 events for Sanofi (risk difference 0.12 (95%CI −0.016 to 0.26)) In the 75+ cohort, 3 and 9 pericarditis events within 4 weeks after vaccination for Pfizer BA.4-5 and Sanofi recipients, respectively, with a cumulative incidence per 10,000 people of 0.089 (95%CI 0.029 to 0.28) for those receiving Pfizer BA.4-5 and 0.27 (0.14 to 0.51) for Sanofi (risk difference 0.18 (95%CI −0.023 to 0.38)). For myocarditis, there were 0 events for Pfizer BA.4/5 and 9 events for Sanofi (risk difference 0.27 (95%CI 0.093 to 0.44)).

### Subgroup analyses

Findings in the main analyses were similar when examining subgroups (age band in the CV group only (50-64, 65-75, 75-79, 80-84, 85+); clinical vulnerability in the 75+ group only; prior COVID-19 vaccine count), although estimated with reduced precision. Supplementary materials provide cumulative incidence plots by subgroup (Figures S3), the corresponding cumulative risk differences, cumulative risk ratios, and period-specific IRRs (Figures S4), and subgroup-specific heterogeneity tests for risk differences and risk ratios (Table S2).

## Discussion

This observational study of COVID-19 vaccine recipients during the spring 2023 booster programme in England is, to our knowledge, the first to directly compare the Pfizer BA.4-5 and Sanofi vaccines for safety and effectiveness against severe COVID-19 outcomes. We estimated that, in both CV (N=492,642 people) and 75+ (N=673,926 people) cohorts, COVID-19 related hospital admissions were less frequent in those receiving the Pfizer BA.4-5 vaccine compared with Sanofi during the first 16 weeks after vaccination. For the more severe COVID-19 outcomes of critical care admission and death, there were very few events resulting in reduced power to identify differences between vaccines.

### Strengths and limitations

The OpenSAFELY-TPP database covers around 45% of the English population and contains rich clinical information which enabled us to closely match Pfizer BA.4-5 and Sanofi recipients to control for potential confounding and compare safety and effectiveness.

To make fair comparisons between the two vaccine types, we exploited the concurrent roll-out of both vaccines across the same eligible population during the same time period. We used 1-1 matching to control for potential differences in the distribution of prognostic factors between the two vaccine groups, and although this led to the omission of a substantial number of people from the final analysis, characteristics were very well-balanced after matching.

As an observational study, we cannot rule out residual confounding, though this is less problematic in comparative effectiveness studies than in studies comparing vaccinated with unvaccinated people, as all people entering the study have sought and received a booster vaccine.

We excluded groups who were unlikely to undergo boosting such as those in palliative care. We also excluded those identified as health and social care workers due to these groups being much more likely to receive their vaccines at their place of work, rather than at walk-in or bookable community vaccination centres, which may introduce bias. For example, occupation type for health care workers may influence both vaccine administration setting (and therefore vaccine type) and risk of infection.

This study does not include anyone who received the Moderna mRNA-1273 vaccine which was also available during the spring 2023 booster programme but was administered much more infrequently.

### Findings in context

Out-of-trial evidence of the effectiveness of Sanofi and Pfizer BA.4-5 vaccines against hospitalisation amongst adults aged 75 years and older in England from a test-negative case control study involving 14,174 eligible tests found slightly higher effectiveness (albeit with low precision) of Pfizer BA.4-5 compared to Sanofi at 10+ weeks, 37.8 (95% C.I: 13.1 to 55.5) and 17.7 (−3.5 to 34.6) respectively, with effectiveness against hospitalisation highest in the period 9 to 13 days post vaccination for both manufacturers (Sanofi: 43.6% (95% C.I.; 20.1 to 60.2%) and Pfizer BA.4-5: 56.4% (95% C.I; 25.8 to 74.4%).(7). However, this study estimated effectiveness of vaccination versus no vaccination in Pfizer BA.4-5 and Sanofi independently, unlike in the present study where direct comparisons were made by carefully balancing characteristics across groups. We are aware of no other planned or published randomised trials making direct endpoint comparisons of boosting with Pfizer BA.4-5 and Sanofi.

## Conclusions

During the spring 2023 COVID-19 booster programme in England, COVID-19-related hospital admissions were slightly more frequent following receipt of the Sanofi vaccine than the Pfizer BA.4-5 vaccine after matching on a number of important potential confounders. These findings were broadly consistent across subgroups, though estimated with less precision.

### Contributions

Conceptualization: WJH, EPKP, VW, TP, BG, MAH, and JACS. Data curation: CDA, EP, WJH, BS, HJC, AW, AM, BM, RMS, CW, ALS, ACAG, and SCJB. Formal analysis: WJH, CDA, and VW. Funding acquisition: BG, and JACS. Methodology: CDA, EP, WJH, EPKP, VW, TP, ALS, MAH, and JACS. Project administration: WJH, CDA, AM, BG, ACAG, and JACS. Resources: WJH, CDA, HJC, AW, AM, RMS, ACAG, BM, BS, SCJB, and BG. Software: WJH, CDA, EP, EPKP, RMS, BS, and SCJB. Supervision: WJH and JACS. Validation: CDA, EP, WJH, EPKP, ALS, RMS, CW and VW. Visualization: WJH, CDA, and EPKP. Writing - original draft: WJH. Writing - review & editing: WJH, CDA, EP, EPKP, VW, TP, HJC, AW, AM, BM, SCJB, BG, MAH, and JACS.

### Information governance and ethical approval

NHS England is the data controller for the NHS England OpenSAFELY COVID-19 Service; TPP is the data processor; all study authors using OpenSAFELY have the approval of NHS England (8). This implementation of OpenSAFELY is hosted within the TPP environment which is accredited to the ISO 27001 information security standard and is NHS IG Toolkit compliant (9).

Patient data has been pseudonymised for analysis and linkage using industry standard cryptographic hashing techniques; all pseudonymised datasets transmitted for linkage onto OpenSAFELY are encrypted; access to the NHS England OpenSAFELY COVID-19 service is via a virtual private network (VPN) connection; the researchers hold contracts with NHS England and only access the platform to initiate database queries and statistical models; all database activity is logged; only aggregate statistical outputs leave the platform environment following best practice for anonymisation of results such as statistical disclosure control for low cell counts (10).

The service adheres to the obligations of the UK General Data Protection Regulation (UK GDPR) and the Data Protection Act 2018. The service previously operated under notices initially issued in February 2020 by the Secretary of State under Regulation 3(4) of the Health Service (Control of Patient Information) Regulations 2002 (COPI Regulations), which required organisations to process confidential patient information for COVID-19 purposes; this set aside the requirement for patient consent (11). As of 1 July 2023, the Secretary of State has requested that NHS England continue to operate the Service under the COVID-19 Directions 2020 (12). In some cases of data sharing, the common law duty of confidence is met using, for example, patient consent or support from the Health Research Authority Confidentiality Advisory Group (13).

Taken together, these provide the legal bases to link patient datasets using the service. GP practices, which provide access to the primary care data, are required to share relevant health information to support the public health response to the pandemic, and have been informed of how the service operates.

This study was approved by the Health Research Authority (REC reference 20/LO/0651) and by London School of Hygiene and Tropical Medicine Ethics Board (reference 21863).

### Data access and verification

Access to the underlying identifiable and potentially re-identifiable pseudonymised electronic health record data is tightly governed by various legislative and regulatory frameworks, and restricted by best practice. The data in the NHS England OpenSAFELY COVID-19 service is drawn from General Practice data across England where TPP is the data processor.

TPP developers initiate an automated process to create pseudonymised records in the core OpenSAFELY database, which are copies of key structured data tables in the identifiable records. These pseudonymised records are linked onto key external data resources that have also been pseudonymised via SHA-512 one-way hashing of NHS numbers using a shared salt. University of Oxford, Bennett Institute for Applied Data Science developers and PIs, who hold contracts with NHS England, have access to the OpenSAFELY pseudonymised data tables to develop the OpenSAFELY tools.

These tools in turn enable researchers with OpenSAFELY data access agreements to write and execute code for data management and data analysis without direct access to the underlying raw pseudonymised patient data, and to review the outputs of this code. All code for the full data management pipeline — from raw data to completed results for this analysis — and for the OpenSAFELY platform as a whole is available for review at www.github.com/OpenSAFELY.

## Patient and Public Involvement and Engagement (PPIE)

OpenSAFELY has developed a publicly available website https://www.opensafely.org/ through which they invite any patient or member of the public to make contact regarding the broader OpenSAFELY project.

## Data Availability

Access to the underlying identifiable and potentially re-identifiable pseudonymised electronic health record data is tightly governed by various legislative and regulatory frameworks, and restricted by best practice. The data in the NHS England OpenSAFELY COVID-19 service is drawn from General Practice data across England where TPP is the data processor. TPP developers initiate an automated process to create pseudonymised records in the core OpenSAFELY database, which are copies of key structured data tables in the identifiable records. These pseudonymised records are linked onto key external data resources that have also been pseudonymised via SHA-512 one-way hashing of NHS numbers using a shared salt. University of Oxford, Bennett Institute for Applied Data Science developers and PIs, who hold contracts with NHS England, have access to the OpenSAFELY pseudonymised data tables to develop the OpenSAFELY tools. These tools in turn enable researchers with OpenSAFELY data access agreements to write and execute code for data management and data analysis without direct access to the underlying raw pseudonymised patient data, and to review the outputs of this code. All code for the full data management pipeline - from raw data to completed results for this analysis - and for the OpenSAFELY platform as a whole is available for review at www.github.com/OpenSAFELY.

## Acknowledgements

We are very grateful for all the support received from the TPP Technical Operations team throughout this work, and for generous assistance from the information governance and database teams at NHS England and the NHS England Transformation Directorate.

