## Supplemental materials for "Comparative safety and effectiveness of Pfizer BA.4-5 versus Sanofi during the spring 2023 COVID-19 booster vaccination programme in England: a matched cohort study in OpenSAFELY-TPP"

#### Figure S1a: Cumulative numbers included in the matched sample for the CV cohort

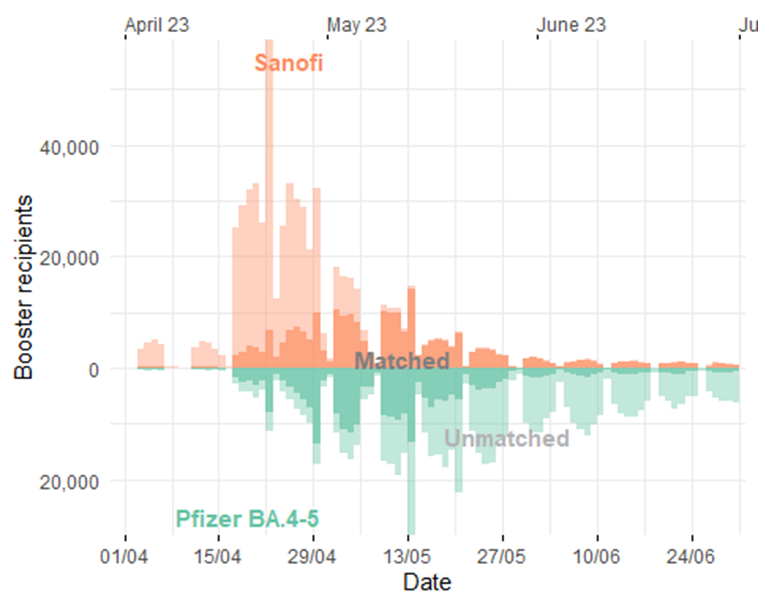

#### Figure S1b: Cumulative numbers included in the matched sample for the 75+ cohort

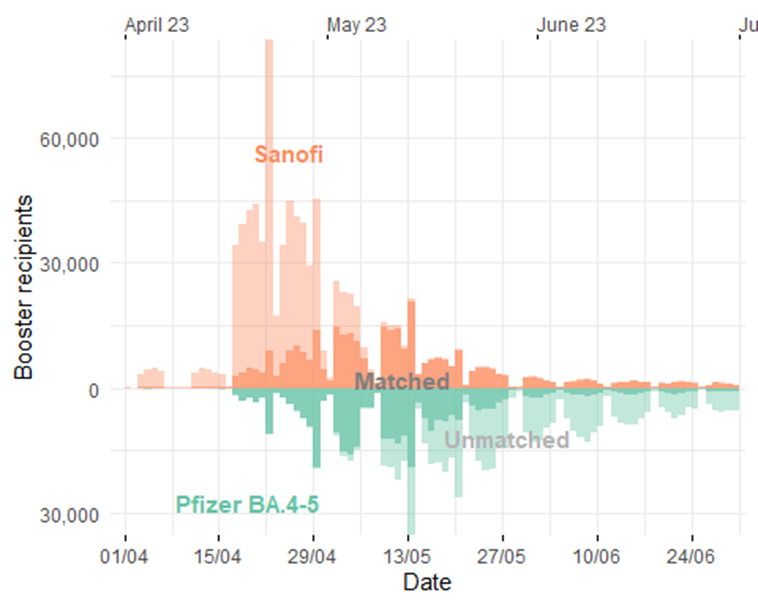

#### Table S1a: Baseline characteristics in the CV cohort between Pfizer BA.4-5 and Sanofi groups, before and after matching

| Variable |  | Before matching | | | After matching | | |
| --- | --- | --- | --- | --- | --- | --- | --- |
|  |  | Pfizer BA.4-5 (N=679311) | Sanofi (N=647241) | SMD | Pfizer BA.4-5 (N=246321) | Sanofi (N=246321) | SMD |
| Previous vaccine count | 2-4 | 164331 (24.2) | 87033 (13.4) | -0.278 | 33447 (13.6) | 33447 (13.6) | - |
|  | 5 | 478329 (70.4) | 541551 (83.7) | 0.319 | 208401 (84.6) | 208401 (84.6) | - |
|  | 6+ | 36657 (5.4) | 18657 (2.9) | -0.126 | 4473 (1.8) | 4473 (1.8) | - |
| Age band | 50-64 | 72945 (10.7) | 1743 (0.3) | -0.472 | 957 (0.4) | 957 (0.4) | - |
|  | 65-74 | 90759 (13.4) | 11253 (1.7) | -0.451 | 6573 (2.7) | 6573 (2.7) | - |
|  | 75-79 | 226779 (33.4) | 233781 (36.1) | 0.057 | 100449 (40.8) | 100449 (40.8) | - |
|  | 80-84 | 149625 (22.0) | 186105 (28.8) | 0.155 | 71127 (28.9) | 71127 (28.9) | - |
|  | 85+ | 139197 (20.5) | 214359 (33.1) | 0.288 | 67209 (27.3) | 67209 (27.3) | - |
| Sex | female | 356025 (52.4) | 332703 (51.4) | -0.020 | 126273 (51.3) | 126273 (51.3) | - |
|  | male | 323289 (47.6) | 314541 (48.6) | 0.020 | 120045 (48.7) | 120045 (48.7) | - |
| Ethnicity | White | 571521 (84.1) | 547221 (84.5) | 0.011 | 204987 (83.2) | 205227 (83.3) | 0.003 |
|  | Black | 3693 (0.5) | 1911 (0.3) | -0.038 | 681 (0.3) | 705 (0.3) | 0.002 |
|  | South Asian | 9855 (1.5) | 6345 (1.0) | -0.043 | 2835 (1.2) | 2643 (1.1) | -0.007 |
|  | Mixed | 1689 (0.2) | 1119 (0.2) | -0.017 | 369 (0.1) | 417 (0.2) | 0.005 |
|  | Other | 3021 (0.4) | 2037 (0.3) | -0.021 | 759 (0.3) | 729 (0.3) | -0.002 |
|  | Unknown | 89529 (13.2) | 88605 (13.7) | 0.015 | 36687 (14.9) | 36603 (14.9) | -0.001 |
| Deprivation | 1 most deprived | 76137 (11.2) | 72573 (11.2) | 0.000 | 23913 (9.7) | 25893 (10.5) | 0.027 |
|  | 2 | 112833 (16.6) | 103299 (16.0) | -0.018 | 38739 (15.7) | 39267 (15.9) | 0.006 |
|  | 3 | 159987 (23.6) | 148365 (22.9) | -0.015 | 58095 (23.6) | 56961 (23.1) | -0.011 |
|  | 4 | 163941 (24.1) | 160653 (24.8) | 0.016 | 61323 (24.9) | 62289 (25.3) | 0.009 |
|  | 5 least deprived | 166419 (24.5) | 162351 (25.1) | 0.014 | 64245 (26.1) | 61911 (25.1) | -0.022 |
| Region | North East and Yorkshire | 103623 (15.3) | 130257 (20.1) | 0.128 | 36693 (14.9) | 36693 (14.9) | - |
|  | North West | 71163 (10.5) | 50679 (7.8) | -0.092 | 20007 (8.1) | 20007 (8.1) | - |
|  | Midlands | 146541 (21.6) | 127437 (19.7) | -0.047 | 57285 (23.3) | 57285 (23.3) | - |
|  | East of England | 164685 (24.2) | 147483 (22.8) | -0.034 | 59463 (24.1) | 59463 (24.1) | - |
|  | London | 15309 (2.3) | 13953 (2.2) | -0.007 | 4107 (1.7) | 4107 (1.7) | - |
|  | South East | 52443 (7.7) | 50235 (7.8) | 0.002 | 19401 (7.9) | 19401 (7.9) | - |
|  | South West | 125553 (18.5) | 127191 (19.7) | 0.030 | 49365 (20.0) | 49365 (20.0) | - |
| Clinically at-risk |  | 679311 (100.0) | 647241 (100.0) | - | 246321 (100.0) | 246321 (100.0) | - |
| Clinically housebound |  | 42165 (6.2) | 39123 (6.0) | -0.007 | 16353 (6.6) | 15819 (6.4) | -0.009 |
| Care/nursing home resident |  | 18195 (2.7) | 55515 (8.6) | 0.258 | 7239 (2.9) | 10329 (4.2) | 0.068 |
| Body Mass Index > 40 kg/m^2 |  | 21627 (3.2) | 12705 (2.0) | -0.077 | 5613 (2.3) | 5535 (2.2) | -0.002 |
| Chronic heart disease |  | 337137 (49.6) | 361659 (55.9) | 0.125 | 137961 (56.0) | 137211 (55.7) | -0.006 |
| Chronic kidney disease |  | 200913 (29.6) | 228813 (35.4) | 0.124 | 85413 (34.7) | 86235 (35.0) | 0.007 |
| Diabetes |  | 174489 (25.7) | 165489 (25.6) | -0.003 | 66915 (27.2) | 66705 (27.1) | -0.002 |
| Chronic liver disease |  | 35679 (5.3) | 25473 (3.9) | -0.063 | 10287 (4.2) | 10419 (4.2) | 0.003 |
| Chronic respiratory disease |  | 135663 (20.0) | 119529 (18.5) | -0.038 | 47139 (19.1) | 46605 (18.9) | -0.006 |
| Asthma |  | 39921 (5.9) | 29547 (4.6) | -0.059 | 12021 (4.9) | 11667 (4.7) | -0.007 |
| Chronic neurological disease |  | 160821 (23.7) | 183603 (28.4) | 0.107 | 63885 (25.9) | 64737 (26.3) | 0.008 |
| Immunosuppressed |  | 141057 (20.8) | 60945 (9.4) | -0.321 | 20349 (8.3) | 20349 (8.3) | - |
| Immunosuppressed (all) |  | 214929 (31.6) | 127869 (19.8) | -0.274 | 45879 (18.6) | 45183 (18.3) | -0.007 |
| Asplenia or poor spleen function |  | 11445 (1.7) | 8739 (1.4) | -0.027 | 3075 (1.2) | 3087 (1.3) | 0.000 |
| Solid organ transplant |  | 6573 (1.0) | 1119 (0.2) | -0.106 | 495 (0.2) | 393 (0.2) | -0.010 |
| HIV/AIDS |  | 435 (0.1) | 57 (0.0) | -0.029 | 15 (0.0) | 21 (0.0) | 0.003 |
| Morbidity count | 0 | 44553 (6.6) | 14463 (2.2) | -0.212 | 4449 (1.8) | 4449 (1.8) | - |
|  | 1 | 293313 (43.2) | 283113 (43.7) | 0.011 | 110085 (44.7) | 110085 (44.7) | - |
|  | 2+ | 341451 (50.3) | 349665 (54.0) | 0.075 | 131787 (53.5) | 131787 (53.5) | - |
| Learning disabilities |  | 3651 (0.5) | 2019 (0.3) | -0.035 | 477 (0.2) | 663 (0.3) | 0.016 |
| Serious mental illness |  | 7365 (1.1) | 6627 (1.0) | -0.006 | 1845 (0.7) | 2061 (0.8) | 0.010 |
| Number of SARS-CoV-2 tests | 0 | 656241 (96.6) | 614091 (94.9) | -0.086 | 239223 (97.1) | 238767 (96.9) | -0.011 |
|  | 1 | 14937 (2.2) | 18471 (2.9) | 0.042 | 4395 (1.8) | 4689 (1.9) | 0.009 |
|  | 2 | 4323 (0.6) | 7071 (1.1) | 0.049 | 1443 (0.6) | 1551 (0.6) | 0.006 |
|  | 3+ | 3807 (0.6) | 7605 (1.2) | 0.066 | 1257 (0.5) | 1311 (0.5) | 0.003 |
| Prior documented SARS-CoV-2 infection |  | 142089 (20.9) | 126675 (19.6) | -0.033 | 40131 (16.3) | 40131 (16.3) | - |
| Previously received Pfizer (original) |  | 647805 (95.4) | 631917 (97.6) | 0.124 | 239379 (97.2) | 240357 (97.6) | 0.025 |
| Previously received AZ |  | 326211 (48.0) | 258753 (40.0) | -0.163 | 103023 (41.8) | 102501 (41.6) | -0.004 |
| Previously received Moderna |  | 374733 (55.2) | 375615 (58.0) | 0.058 | 143805 (58.4) | 146181 (59.3) | 0.020 |
| Previously received Pfizer/BA.1 |  | 273369 (40.2) | 182409 (28.2) | -0.256 | 84423 (34.3) | 78921 (32.0) | -0.047 |
| Previously received Pfizer/XBB.1.5 |  | 0 (0.0) | 0 (0.0) | - | 0 (0.0) | 0 (0.0) | - |
| Previously received Moderna/Omicron |  | 349773 (51.5) | 406845 (62.9) | 0.231 | 146367 (59.4) | 149811 (60.8) | 0.029 |
| Previously received Moderna/XBB.1.5 |  | 3 (0.0) | 0 (0.0) | -0.003 | 0 (0.0) | 0 (0.0) | - |

For variables that were matched exactly the SMD is zero by design, and so are not shown.

#### Table S1b: Baseline characteristics in the 75+ cohort between Pfizer BA.4-5 and Sanofi groups, before and after matching

| Variable |  | Before matching | | | After matching | | |
| --- | --- | --- | --- | --- | --- | --- | --- |
|  |  | Pfizer BA.4-5 (N=724089) | Sanofi (N=876171) | SMD | Pfizer BA.4-5 (N=336963) | Sanofi (N=336963) | SMD |
| Previous vaccine count | 2-4 | 130365 (18.0) | 118881 (13.6) | -0.122 | 46917 (13.9) | 46917 (13.9) | - |
|  | 5 | 579297 (80.0) | 737319 (84.2) | 0.108 | 285909 (84.8) | 285909 (84.8) | - |
|  | 6+ | 14427 (2.0) | 19971 (2.3) | 0.020 | 4137 (1.2) | 4137 (1.2) | - |
| Age band | 75-79 | 354609 (49.0) | 364149 (41.6) | -0.149 | 158991 (47.2) | 158991 (47.2) | - |
|  | 80-84 | 202215 (27.9) | 253599 (28.9) | 0.023 | 97071 (28.8) | 97071 (28.8) | - |
|  | 85+ | 167259 (23.1) | 258423 (29.5) | 0.146 | 80901 (24.0) | 80901 (24.0) | - |
| Sex | female | 396483 (54.8) | 475533 (54.3) | -0.010 | 183321 (54.4) | 183321 (54.4) | - |
|  | male | 327603 (45.2) | 400641 (45.7) | 0.010 | 153639 (45.6) | 153639 (45.6) | - |
| Ethnicity | White | 609795 (84.2) | 743691 (84.9) | 0.018 | 282315 (83.8) | 281973 (83.7) | -0.003 |
|  | Black | 2481 (0.3) | 2205 (0.3) | -0.017 | 801 (0.2) | 825 (0.2) | 0.001 |
|  | South Asian | 8307 (1.1) | 7713 (0.9) | -0.027 | 3393 (1.0) | 3231 (1.0) | -0.005 |
|  | Mixed | 1263 (0.2) | 1383 (0.2) | -0.004 | 471 (0.1) | 531 (0.2) | 0.005 |
|  | Other | 2751 (0.4) | 2787 (0.3) | -0.010 | 1017 (0.3) | 1005 (0.3) | -0.001 |
|  | Unknown | 99495 (13.7) | 118389 (13.5) | -0.007 | 48969 (14.5) | 49395 (14.7) | 0.004 |
| Deprivation | 1 most deprived | 71301 (9.8) | 88959 (10.2) | 0.010 | 29361 (8.7) | 32259 (9.6) | 0.030 |
|  | 2 | 114135 (15.8) | 134199 (15.3) | -0.012 | 50811 (15.1) | 51561 (15.3) | 0.006 |
|  | 3 | 170745 (23.6) | 200247 (22.9) | -0.017 | 79131 (23.5) | 77541 (23.0) | -0.011 |
|  | 4 | 179601 (24.8) | 222099 (25.3) | 0.013 | 85317 (25.3) | 86823 (25.8) | 0.010 |
|  | 5 least deprived | 188301 (26.0) | 230661 (26.3) | 0.007 | 92337 (27.4) | 88779 (26.3) | -0.024 |
| Region | North East and Yorkshire | 105831 (14.6) | 171405 (19.6) | 0.132 | 47919 (14.2) | 47919 (14.2) | - |
|  | North West | 79365 (11.0) | 65373 (7.5) | -0.121 | 26811 (8.0) | 26811 (8.0) | - |
|  | Midlands | 154251 (21.3) | 173151 (19.8) | -0.038 | 77637 (23.0) | 77637 (23.0) | - |
|  | East of England | 180267 (24.9) | 203181 (23.2) | -0.040 | 83025 (24.6) | 83025 (24.6) | - |
|  | London | 14067 (1.9) | 19227 (2.2) | 0.018 | 5823 (1.7) | 5823 (1.7) | - |
|  | South East | 57903 (8.0) | 69459 (7.9) | -0.003 | 27747 (8.2) | 27747 (8.2) | - |
|  | South West | 132405 (18.3) | 174369 (19.9) | 0.041 | 67995 (20.2) | 67995 (20.2) | - |
| Clinically at-risk |  | 515607 (71.2) | 634245 (72.4) | 0.026 | 238791 (70.9) | 238791 (70.9) | - |
| Clinically housebound |  | 43017 (5.9) | 42213 (4.8) | -0.050 | 17727 (5.3) | 17073 (5.1) | -0.009 |
| Care/nursing home resident |  | 14085 (1.9) | 52665 (6.0) | 0.209 | 7137 (2.1) | 8961 (2.7) | 0.035 |
| Body Mass Index > 40 kg/m^2 |  | 14469 (2.0) | 14295 (1.6) | -0.027 | 5847 (1.7) | 5769 (1.7) | -0.002 |
| Chronic heart disease |  | 289917 (40.0) | 357159 (40.8) | 0.015 | 135303 (40.2) | 134541 (39.9) | -0.005 |
| Chronic kidney disease |  | 178407 (24.6) | 226917 (25.9) | 0.029 | 84159 (25.0) | 85065 (25.2) | 0.006 |
| Diabetes |  | 139281 (19.2) | 162231 (18.5) | -0.018 | 65043 (19.3) | 64863 (19.2) | -0.001 |
| Chronic liver disease |  | 20961 (2.9) | 24507 (2.8) | -0.006 | 9573 (2.8) | 9879 (2.9) | 0.005 |
| Chronic respiratory disease |  | 99963 (13.8) | 117021 (13.4) | -0.013 | 45231 (13.4) | 44997 (13.4) | -0.002 |
| Asthma |  | 24801 (3.4) | 28767 (3.3) | -0.008 | 11313 (3.4) | 11121 (3.3) | -0.003 |
| Chronic neurological disease |  | 135807 (18.8) | 177981 (20.3) | 0.039 | 62499 (18.5) | 62355 (18.5) | -0.001 |
| Immunosuppressed |  | 44007 (6.1) | 56379 (6.4) | 0.015 | 16845 (5.0) | 16845 (5.0) | - |
| Immunosuppressed (all) |  | 96309 (13.3) | 122259 (14.0) | 0.019 | 41205 (12.2) | 40989 (12.2) | -0.002 |
| Asplenia or poor spleen function |  | 6753 (0.9) | 8463 (1.0) | 0.003 | 2895 (0.9) | 2913 (0.9) | 0.001 |
| Solid organ transplant |  | 717 (0.1) | 921 (0.1) | 0.002 | 285 (0.1) | 261 (0.1) | -0.003 |
| HIV/AIDS |  | 33 (0.0) | 39 (0.0) | 0.000 | 9 (0.0) | 9 (0.0) | 0.000 |
| Morbidity count | 0 | 216249 (29.9) | 252543 (28.8) | -0.023 | 100983 (30.0) | 100983 (30.0) | - |
|  | 1 | 230385 (31.8) | 280227 (32.0) | 0.004 | 107685 (32.0) | 107685 (32.0) | - |
|  | 2+ | 277455 (38.3) | 343401 (39.2) | 0.018 | 128301 (38.1) | 128301 (38.1) | - |
| Learning disabilities |  | 945 (0.1) | 1101 (0.1) | -0.001 | 333 (0.1) | 327 (0.1) | -0.001 |
| Serious mental illness |  | 4443 (0.6) | 5547 (0.6) | 0.002 | 1707 (0.5) | 1725 (0.5) | 0.001 |
| Number of SARS-CoV-2 tests | 0 | 709209 (97.9) | 840621 (95.9) | -0.117 | 329265 (97.7) | 328977 (97.6) | -0.006 |
|  | 1 | 9771 (1.3) | 20205 (2.3) | 0.071 | 4899 (1.5) | 5091 (1.5) | 0.005 |
|  | 2 | 2925 (0.4) | 7467 (0.9) | 0.057 | 1521 (0.5) | 1605 (0.5) | 0.004 |
|  | 3+ | 2181 (0.3) | 7881 (0.9) | 0.078 | 1275 (0.4) | 1293 (0.4) | 0.001 |
| Prior documented SARS-CoV-2 infection |  | 109431 (15.1) | 152577 (17.4) | 0.062 | 49149 (14.6) | 49149 (14.6) | - |
| Previously received Pfizer (original) |  | 698523 (96.5) | 855873 (97.7) | 0.072 | 327315 (97.1) | 328821 (97.6) | 0.028 |
| Previously received AZ |  | 323169 (44.6) | 358203 (40.9) | -0.076 | 144291 (42.8) | 143409 (42.6) | -0.005 |
| Previously received Moderna |  | 423813 (58.5) | 509295 (58.1) | -0.008 | 197835 (58.7) | 200649 (59.5) | 0.017 |
| Previously received Pfizer/BA.1 |  | 272277 (37.6) | 244287 (27.9) | -0.208 | 113985 (33.8) | 106809 (31.7) | -0.045 |
| Previously received Pfizer/XBB.1.5 |  | 3 (0.0) | 3 (0.0) | 0.000 | 0 (0.0) | 3 (0.0) | 0.004 |
| Previously received Moderna/Omicron |  | 395895 (54.7) | 557307 (63.6) | 0.182 | 202431 (60.1) | 206919 (61.4) | 0.027 |
| Previously received Moderna/XBB.1.5 |  | 3 (0.0) | 3 (0.0) | 0.000 | 0 (0.0) | 0 (0.0) | NA |

For variables that were matched exactly the SMD is zero by design, and so are not shown.

#### Figure S2a: Standardised Mean Differences for CV cohort between Pfizer BA.4-5 and Sanofi groups, before and after matching

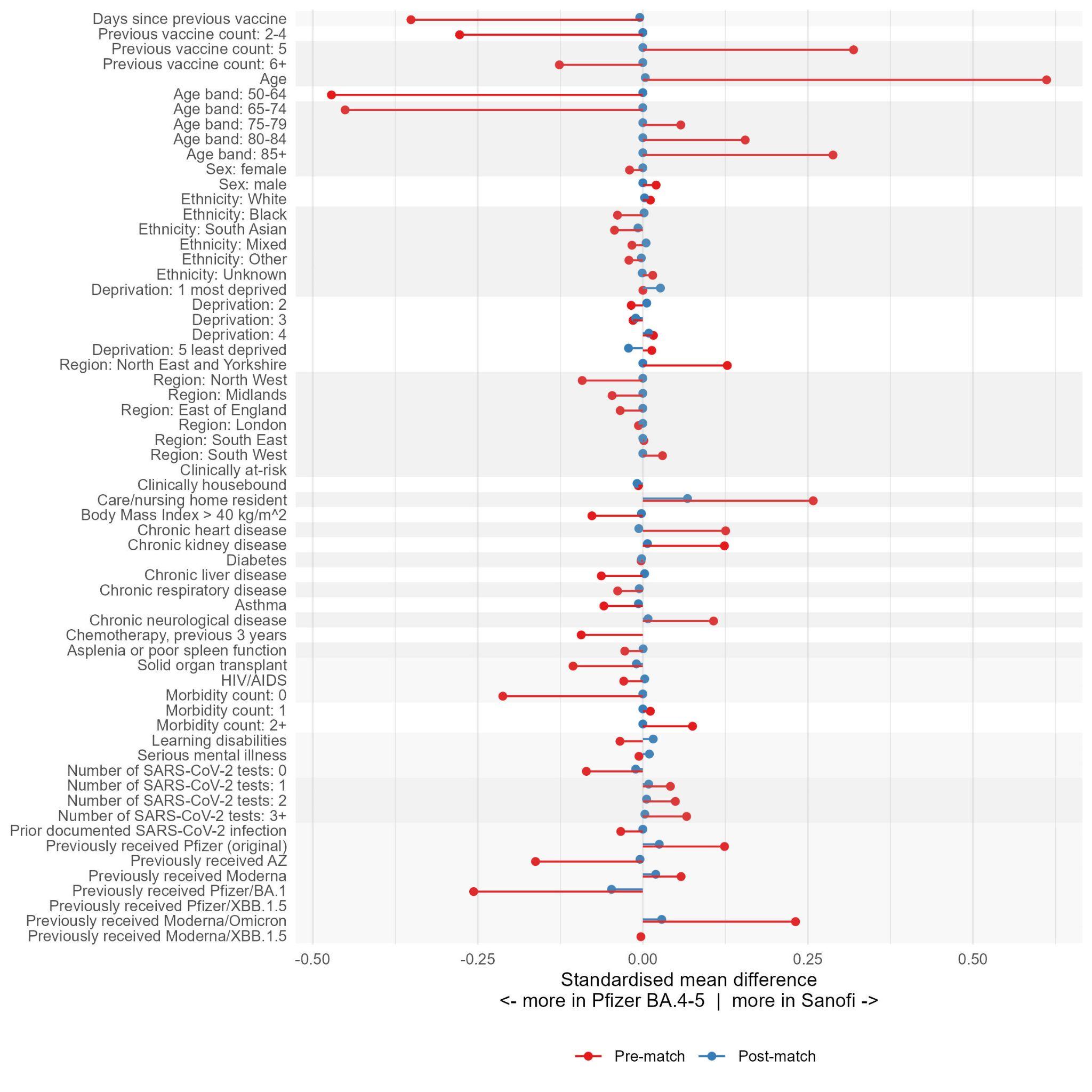

#### Figure S2b: Standardised Mean Differences for 75+ cohort between Pfizer BA.4-5 and Sanofi groups, before and after matching

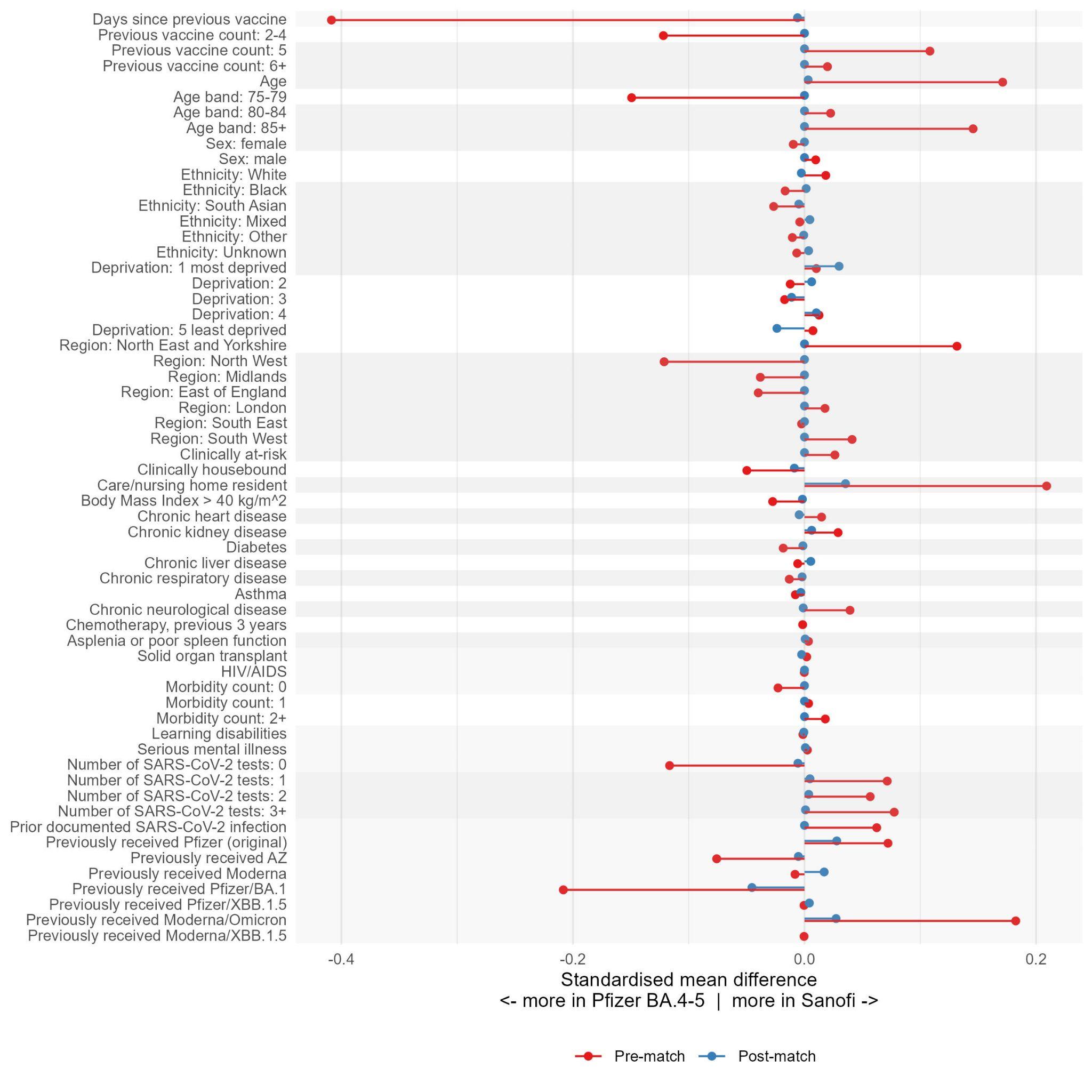

#### Table S2a: Risk differences, risk ratios, and incidence rate ratios in the CV cohort, with p-values for subgroup heterogeneity

| Sub-group |  | Risk difference per 1,000 people (95% CI) | P- value | Risk ratio (95% CI) | P- value | IRR (95% CI) | P- value |
| --- | --- | --- | --- | --- | --- | --- | --- |
| COVID-19 hospitalisation | | | | | | | |
| Main |  | 0.42 (0.13 to 0.71) |  | 1.19 (1.05-1.34) |  | 1.19 (1.06-1.34) |  |
| Age | 50-64 | 3.18 (−0.41 to 6.78) | 0.424 | Inf ( - ) |  | Inf ( - ) |  |
|  | 65-74 | −0.020 (−1.32 to 1.28) |  | 0.99 (0.39-2.48) |  | 1.01 (0.40-2.54) |  |
|  | 75-79 | 0.26 (−0.10 to 0.62) |  | 1.18 (0.93-1.50) |  | 1.18 (0.94-1.50) |  |
|  | 80-84 | 0.54 (0.047 to 1.04) |  | 1.30 (1.02-1.66) |  | 1.30 (1.03-1.66) |  |
|  | 85+ | 0.59 (−0.15 to 1.33) |  | 1.15 (0.97-1.37) |  | 1.15 (0.97-1.36) |  |
| Prior COVID-19 vaccine count | 2-4 | 0.25 (−0.47 to 0.98) | 0.589 | 1.13 (0.79-1.63) | 0.650 | 1.10 (0.77-1.58) | 0.666 |
|  | 5 | 0.40 (0.084 to 0.72) |  | 1.18 (1.03-1.34) |  | 1.19 (1.05-1.35) |  |
|  | 6+ | 1.47 (−0.74 to 3.67) |  | 1.72 (0.76-3.94) |  | 1.67 (0.73-3.81) |  |
| COVID-19 critical care | | | | | | | |
| Main |  | 0.083 (0.040 to 0.13) |  | 7.80 (2.32-26.16) |  | 6.99 (2.08-23.43) |  |
| Age | 50-64 | 0.000 (0.000 to 0.000) |  | - ( - ) |  | - ( - ) |  |
|  | 65-74 | 0.000 (0.000 to 0.000) |  | - ( - ) |  | - ( - ) |  |
|  | 75-79 | 0.070 (−0.004 to 0.14) |  | 3.31 (0.90-12.25) |  | 2.99 (0.81-11.06) |  |
|  | 80-84 | −0.005 (−0.078 to 0.067) |  | 0.89 (0.18-4.42) |  | 1.00 (0.20-4.94) |  |
|  | 85+ | 0.000 (−0.072 to 0.072) |  | 1.00 (0.20-4.95) |  | 1.00 (0.20-4.95) |  |
| Prior COVID-19 vaccine count | 2-4 | 0.10 (−0.013 to 0.22) | 0.244 | Inf ( - ) |  | Inf ( - ) |  |
|  | 5 | 0.063 (0.020 to 0.11) |  | 5.33 (1.54-18.43) |  | 4.99 (1.44-17.24) |  |
|  | 6+ | 0.69 (−0.090 to 1.47) |  | Inf ( - ) |  | Inf ( - ) |  |
| COVID-19 death | | | | | | | |
| Main |  | 0.004 (−0.069 to 0.078) |  | 1.03 (0.63-1.68) |  | 1.00 (0.62-1.62) |  |
| Age | 50-64 | 0.000 (0.000 to 0.000) |  | - ( - ) |  | - ( - ) |  |
|  | 65-74 | 0.46 (−0.061 to 0.99) |  | Inf ( - ) |  | Inf ( - ) |  |
|  | 75-79 | 0.081 (0.000 to 0.16) |  | 3.69 (1.00-13.68) |  | 2.99 (0.81-11.06) |  |
|  | 80-84 | 0.088 (−0.010 to 0.19) |  | 3.09 (0.84-11.40) |  | 2.99 (0.81-11.06) |  |
|  | 85+ | −0.062 (−0.26 to 0.13) |  | 0.81 (0.41-1.58) |  | 0.71 (0.37-1.38) |  |
| Prior COVID-19 vaccine count | 2-4 | 0.000 (−0.14 to 0.15) | 0.114 | 1.01 (0.20-4.98) |  | 1.00 (0.20-4.95) |  |
|  | 5 | −0.075 (−0.15 to 0.002) |  | 0.58 (0.34-1.01) |  | 0.64 (0.37-1.10) |  |
|  | 6+ | 0.67 (−0.088 to 1.43) |  | Inf ( - ) |  | Inf ( - ) |  |
| Non-COVID-19 death | | | | | | | |
| Main |  | 0.33 (−0.25 to 0.91) |  | 1.03 (0.98-1.09) |  | 1.04 (0.98-1.10) |  |
| Age | 50-64 | 8.26 (−0.006 to 16.5) | 0.006 | 3.63 (0.98-13.38) | 0.009 | 3.01 (0.82-11.13) | 0.016 |
|  | 65-74 | 5.83 (2.37 to 9.29) |  | 1.83 (1.27-2.63) |  | 1.81 (1.26-2.61) |  |
|  | 75-79 | −0.056 (−0.70 to 0.59) |  | 0.99 (0.87-1.12) |  | 1.00 (0.88-1.13) |  |
|  | 80-84 | 0.31 (−0.64 to 1.26) |  | 1.04 (0.92-1.17) |  | 1.04 (0.93-1.18) |  |
|  | 85+ | 0.37 (−1.19 to 1.92) |  | 1.02 (0.94-1.10) |  | 1.02 (0.94-1.10) |  |
| Prior COVID-19 vaccine count | 2-4 | −0.26 (−1.88 to 1.36) | 0.722 | 0.98 (0.83-1.14) | 0.715 | 0.98 (0.84-1.15) | 0.769 |
|  | 5 | 0.40 (−0.23 to 1.03) |  | 1.04 (0.98-1.11) |  | 1.04 (0.98-1.11) |  |
|  | 6+ | −0.38 (−4.69 to 3.93) |  | 0.96 (0.64-1.46) |  | 1.00 (0.66-1.51) |  |
| Pericarditis | | | | | | | |
| Main |  | 0.024 (−0.003 to 0.052) |  | 3.00 (0.81-11.09) |  | 3.00 (0.81-11.09) |  |
| Age | 50-64 | 0.000 (0.000 to 0.000) |  | - ( - ) |  | - ( - ) |  |
|  | 65-74 | −0.46 (−0.97 to 0.060) |  | 0.0000 ( - ) |  | 0.0000 ( - ) |  |
|  | 75-79 | 0.030 (−0.004 to 0.064) |  | Inf ( - ) |  | Inf ( - ) |  |
|  | 80-84 | 0.000 (−0.068 to 0.067) |  | 1.00 (0.20-4.94 |  | 1.00 (0.20-4.95) |  |
|  | 85+ | 0.045 (−0.006 to 0.096) |  | Inf ( - ) |  | Inf ( - ) |  |
| Prior COVID-19 vaccine count | 2-4 | 0.000 (−0.14 to 0.14) |  | 1.00 (0.20-4.96) |  | 1.00 (0.20-4.96) |  |
|  | 5 | 0.029 (−0.004 to 0.062) |  | 3.00 (0.81-11.08) |  | 3.00 (0.81-11.08) |  |
|  | 6+ | 0.000 (0.000 to 0.000) |  | - ( - ) |  | - ( - ) |  |
| Myocarditis | | | | | | | |
| Main |  | 0.012 (−0.002 to 0.026) |  | Inf ( - ) |  | Inf ( - ) |  |
| Age | 50-64 | 0.000 (0.000 to 0.000) |  | - ( - ) |  | - ( - ) |  |
|  | 65-74 | 0.000 (0.000 to 0.000) |  | - ( - ) |  | - ( - ) |  |
|  | 75-79 | 0.030 (−0.004 to 0.064) |  | Inf ( - ) |  | Inf ( - ) |  |
|  | 80-84 | 0.042 (−0.006 to 0.090) |  | Inf ( - ) |  | Inf ( - ) |  |
|  | 85+ | 0.045 (−0.006 to 0.095) |  | Inf ( - ) |  | Inf ( - ) |  |
| Prior COVID-19 vaccine count | 2-4 | 0.090 (−0.012 to 0.19) |  | Inf ( - ) |  | Inf ( - ) |  |
|  | 5 | 0.014 (−0.002 to 0.031) |  | Inf ( - ) |  | Inf ( - ) |  |
|  | 6+ | 0.000 (0.000 to 0.000) |  | - ( - ) |  | - ( - ) |  |

#### Table S2b: Risk differences, risk ratios, and incidence rate ratios in the age75plus cohort, with p-values for subgroup heterogeneity

| Sub-group |  | Risk difference per 1,000 people (95% CI) | P- value | Risk ratio (95% CI) | P- value | IRR (95% CI) | P- value |
| --- | --- | --- | --- | --- | --- | --- | --- |
| COVID-19 hospitalisation | | | | | | | |
| Main |  | 0.29 (0.073 to 0.51) |  | 1.17 (1.04-1.31) |  | 1.18 (1.05-1.32) |  |
| Age | 75-79 | 0.13 (−0.11 to 0.36) | 0.210 | 1.13 (0.90-1.42) | 0.559 | 1.13 (0.90-1.41) | 0.564 |
|  | 80-84 | 0.47 (0.086 to 0.85) |  | 1.32 (1.05-1.67) |  | 1.32 (1.05-1.66) |  |
|  | 85+ | 0.54 (−0.095 to 1.17) |  | 1.15 (0.98-1.36) |  | 1.16 (0.98-1.37) |  |
| Clinically at-risk | Not clinically at-risk | 0.063 (−0.16 to 0.29) | 0.087 | 1.12 (0.75-1.69) | 0.846 | 1.13 (0.76-1.69) | 0.842 |
|  | Clinically at-risk | 0.39 (0.091 to 0.69) |  | 1.17 (1.04-1.32) |  | 1.18 (1.05-1.33) |  |
| Prior COVID-19 vaccine count | 2-4 | −0.030 (−0.52 to 0.46) | 0.222 | 0.98 (0.68-1.42) | 0.395 | 1.00 (0.69-1.44) | 0.457 |
|  | 5 | 0.35 (0.11 to 0.59) |  | 1.20 (1.06-1.36) |  | 1.20 (1.06-1.36) |  |
|  | 6+ | 1.59 (−0.80 to 3.98) |  | 1.72 (0.75-3.93) |  | 1.67 (0.73-3.81) |  |
| COVID-19 critical care | | | | | | | |
| Main |  | 0.031 (−0.007 to 0.069) |  | 1.83 (0.83-4.05) |  | 2.33 (1.07-5.08) |  |
| Age | 75-79 | 0.044 (−0.003 to 0.090) | 0.196 | 3.30 (0.89-12.21) | 0.391 | 2.99 (0.81-11.05) | 0.507 |
|  | 80-84 | −0.004 (−0.056 to 0.049) |  | 0.90 (0.18-4.45) |  | 1.00 (0.20-4.94) |  |
|  | 85+ | 0.084 (−0.006 to 0.17) |  | 3.23 (0.88-11.95) |  | 3.00 (0.81-11.07) |  |
| Clinically at-risk | Not clinically at-risk | 0.000 (−0.049 to 0.049) | 0.011 | 1.00 (0.20-4.94) | 0.044 | 1.00 (0.20-4.94) | 0.057 |
|  | Clinically at-risk | 0.086 (0.041 to 0.13) |  | 7.81 (2.33-26.19) |  | 6.99 (2.08-23.42) |  |
| Prior COVID-19 vaccine count | 2-4 | 0.072 (−0.009 to 0.15) | 0.208 | Inf ( - ) |  | Inf ( - ) |  |
|  | 5 | 0.038 (−0.007 to 0.083) |  | 1.88 (0.85-4.15) |  | 2.33 (1.07-5.08) |  |
|  | 6+ | 0.75 (−0.098 to 1.59) |  | Inf ( - ) |  | Inf ( - ) |  |
| COVID-19 death | | | | | | | |
| Main |  | 0.001 (−0.050 to 0.053) |  | 1.01 (0.62-1.65) |  | 1.00 (0.62-1.62) |  |
| Age | 75-79 | 0.051 (0.000 to 0.10) | 0.273 | 3.68 (0.99-13.63) | 0.031 | 2.99 (0.81-11.05) | 0.047 |
|  | 80-84 | 0.064 (−0.007 to 0.14) |  | 3.08 (0.83-11.37) |  | 2.99 (0.81-11.05) |  |
|  | 85+ | −0.073 (−0.23 to 0.081) |  | 0.73 (0.38-1.42) |  | 0.71 (0.37-1.38) |  |
| Clinically at-risk | Not clinically at-risk | −0.001 (−0.050 to 0.049) | 0.489 | 0.98 (0.20-4.85) | 0.811 | 1.00 (0.20-4.94) | 0.815 |
|  | Clinically at-risk | −0.031 (−0.10 to 0.039) |  | 0.80 (0.48-1.33) |  | 0.82 (0.49-1.36) |  |
| Prior COVID-19 vaccine count | 2-4 | 0.000 (−0.10 to 0.10) | 0.184 | 1.00 (0.20-4.97) |  | 1.00 (0.20-4.94) |  |
|  | 5 | −0.028 (−0.088 to 0.031) |  | 0.78 (0.47-1.31) |  | 0.82 (0.49-1.36) |  |
|  | 6+ | 0.73 (−0.095 to 1.55) |  | Inf ( - ) |  | Inf ( - ) |  |
| Non-COVID-19 death | | | | | | | |
| Main |  | 0.17 (−0.26 to 0.60) |  | 1.02 (0.97-1.08) |  | 1.02 (0.97-1.08) |  |
| Age | 75-79 | 0.062 (−0.37 to 0.50) | 0.850 | 1.02 (0.90-1.15) | 0.831 | 1.02 (0.90-1.15) | 0.832 |
|  | 80-84 | 0.31 (−0.41 to 1.02) |  | 1.05 (0.93-1.18) |  | 1.05 (0.94-1.18) |  |
|  | 85+ | 0.12 (−1.21 to 1.44) |  | 1.01 (0.93-1.09) |  | 1.01 (0.93-1.09) |  |
| Clinically at-risk | Not clinically at-risk | 0.18 (−0.20 to 0.57) | 0.945 | 1.11 (0.89-1.39) | 0.460 | 1.08 (0.86-1.34) | 0.661 |
|  | Clinically at-risk | 0.21 (−0.38 to 0.80) |  | 1.02 (0.96-1.08) |  | 1.02 (0.96-1.08) |  |
| Prior COVID-19 vaccine count | 2-4 | −0.68 (−1.81 to 0.45) | 0.306 | 0.91 (0.77-1.07) | 0.316 | 0.92 (0.79-1.08) | 0.371 |
|  | 5 | 0.22 (−0.25 to 0.69) |  | 1.03 (0.97-1.09) |  | 1.03 (0.97-1.10) |  |
|  | 6+ | 1.32 (−3.15 to 5.79) |  | 1.13 (0.74-1.74) |  | 1.16 (0.75-1.77) |  |
| Pericarditis | | | | | | | |
| Main |  | 0.018 (−0.002 to 0.038) |  | 3.00 (0.81-11.09) |  | 3.00 (0.81-11.08) |  |
| Age | 75-79 | 0.019 (−0.002 to 0.040) | 0.699 | Inf ( - ) |  | Inf ( - ) |  |
|  | 80-84 | 0.000 (−0.050 to 0.049) |  | 1.00 (0.20-4.95) |  | 1.00 (0.20-4.95) |  |
|  | 85+ | 0.000 (−0.059 to 0.060) |  | 1.00 (0.20-4.97) |  | 1.00 (0.20-4.96) |  |
| Clinically at-risk | Not clinically at-risk | −0.031 (−0.065 to 0.004) | 0.015 | 0.0000 ( - ) |  | 0.0000 ( - ) |  |
|  | Clinically at-risk | 0.025 (−0.003 to 0.054) |  | 3.00 (0.81-11.08) |  | 3.00 (0.81-11.08) |  |
| Prior COVID-19 vaccine count | 2-4 | 0.064 (−0.008 to 0.14) |  | Inf ( - ) |  | Inf ( - ) |  |
|  | 5 | 0.021 (−0.003 to 0.045) |  | 3.00 (0.81-11.09) |  | 3.00 (0.81-11.08) |  |
|  | 6+ | 0.000 (0.000 to 0.000) |  | - ( - ) |  | - ( - ) |  |
| Myocarditis | | | | | | | |
| Main |  | 0.027 (0.009 to 0.044) |  | Inf ( - ) |  | Inf ( - ) |  |
| Age | 75-79 | 0.019 (−0.002 to 0.040) | 0.690 | Inf ( - ) |  | Inf ( - ) |  |
|  | 80-84 | 0.031 (−0.004 to 0.066) |  | Inf ( - ) |  | Inf ( - ) |  |
|  | 85+ | 0.037 (−0.005 to 0.079) |  | Inf ( - ) |  | Inf ( - ) |  |
| Clinically at-risk | Not clinically at-risk | 0.031 (−0.004 to 0.065) | 0.346 | Inf ( - ) |  | Inf ( - ) |  |
|  | Clinically at-risk | 0.013 (−0.002 to 0.027) |  | Inf ( - ) |  | Inf ( - ) |  |
| Prior COVID-19 vaccine count | 2-4 | 0.064 (−0.008 to 0.14) |  | Inf ( - ) |  | Inf ( - ) |  |
|  | 5 | 0.010 (−0.001 to 0.022) |  | Inf ( - ) |  | Inf ( - ) |  |
|  | 6+ | 0.000 (0.000 to 0.000) |  | - ( - ) |  | - ( - ) |  |

#### Table S3a: Summary of follow-up time for CV cohort

| Vaccine | Weeks of follow-up | | | | |
| --- | --- | --- | --- | --- | --- |
|  | Total | Mean | Median | Q1 | Q3 |
| COVID-19 hospitalisation | | | | | |
| pfizer/BA.4-5 | 3 705 753.9 | 15.0 | 16.0 | 15.0 | 16.0 |
| Sanofi | 3 711 210.4 | 15.1 | 16.0 | 15.1 | 16.0 |
| Both | 7 416 964.3 | 15.1 | 16.0 | 15.0 | 16.0 |
| COVID-19 critical care | | | | | |
| pfizer/BA.4-5 | 3 708 523.3 | 15.1 | 16.0 | 15.0 | 16.0 |
| Sanofi | 3 714 624.4 | 15.1 | 16.0 | 15.1 | 16.0 |
| Both | 7 423 147.7 | 15.1 | 16.0 | 15.0 | 16.0 |
| COVID-19 death | | | | | |
| pfizer/BA.4-5 | 3 708 545.6 | 15.1 | 16.0 | 15.0 | 16.0 |
| Sanofi | 3 714 705.0 | 15.1 | 16.0 | 15.1 | 16.0 |
| Both | 7 423 250.6 | 15.1 | 16.0 | 15.0 | 16.0 |
| Non-COVID-19 death | | | | | |
| pfizer/BA.4-5 | 3 708 545.6 | 15.1 | 16.0 | 15.0 | 16.0 |
| Sanofi | 3 714 705.0 | 15.1 | 16.0 | 15.1 | 16.0 |
| Both | 7 423 250.6 | 15.1 | 16.0 | 15.0 | 16.0 |
| Pericarditis | | | | | |
| pfizer/BA.4-5 | 983 527.3 | 4.0 | 4.0 | 4.0 | 4.0 |
| Sanofi | 983 193.0 | 4.0 | 4.0 | 4.0 | 4.0 |
| Both | 1 966 720.3 | 4.0 | 4.0 | 4.0 | 4.0 |
| Myocarditis | | | | | |
| pfizer/BA.4-5 | 983 530.7 | 4.0 | 4.0 | 4.0 | 4.0 |
| Sanofi | 983 194.7 | 4.0 | 4.0 | 4.0 | 4.0 |
| Both | 1 966 725.4 | 4.0 | 4.0 | 4.0 | 4.0 |

#### Table S3b: Summary of follow-up time for 75+ cohort

| Vaccine | Weeks of follow-up | | | | |
| --- | --- | --- | --- | --- | --- |
|  | Total | Mean | Median | Q1 | Q3 |
| COVID-19 hospitalisation | | | | | |
| pfizer/BA.4-5 | 5 078 234.1 | 15.1 | 16.0 | 15.0 | 16.0 |
| Sanofi | 5 088 333.0 | 15.1 | 16.0 | 15.1 | 16.0 |
| Both | 10 166 567.1 | 15.1 | 16.0 | 15.0 | 16.0 |
| COVID-19 critical care | | | | | |
| pfizer/BA.4-5 | 5 081 180.1 | 15.1 | 16.0 | 15.0 | 16.0 |
| Sanofi | 5 091 928.7 | 15.1 | 16.0 | 15.1 | 16.0 |
| Both | 10 173 108.9 | 15.1 | 16.0 | 15.0 | 16.0 |
| COVID-19 death | | | | | |
| pfizer/BA.4-5 | 5 081 213.6 | 15.1 | 16.0 | 15.0 | 16.0 |
| Sanofi | 5 092 016.1 | 15.1 | 16.0 | 15.1 | 16.0 |
| Both | 10 173 229.7 | 15.1 | 16.0 | 15.0 | 16.0 |
| Non-COVID-19 death | | | | | |
| pfizer/BA.4-5 | 5 081 213.6 | 15.1 | 16.0 | 15.0 | 16.0 |
| Sanofi | 5 092 016.1 | 15.1 | 16.0 | 15.1 | 16.0 |
| Both | 10 173 229.7 | 15.1 | 16.0 | 15.0 | 16.0 |
| Pericarditis | | | | | |
| pfizer/BA.4-5 | 1 345 724.1 | 4.0 | 4.0 | 4.0 | 4.0 |
| Sanofi | 1 345 418.1 | 4.0 | 4.0 | 4.0 | 4.0 |
| Both | 2 691 142.3 | 4.0 | 4.0 | 4.0 | 4.0 |
| Myocarditis | | | | | |
| pfizer/BA.4-5 | 1 345 728.4 | 4.0 | 4.0 | 4.0 | 4.0 |
| Sanofi | 1 345 420.7 | 4.0 | 4.0 | 4.0 | 4.0 |
| Both | 2 691 149.1 | 4.0 | 4.0 | 4.0 | 4.0 |

**Table S4a: Testing rates after boosting for the CV cohort**

| Sub-group |  | Pfizer BA.4-5 | | Sanofi | |
| --- | --- | --- | --- | --- | --- |
|  |  | Person-weeks | All tests | Person-weeks | All tests |
| Main |  | 3 945 566 | 0.000 | 3 978 322 | 0.000 |
| Age | 50-64 | 16 321 | 0.001 | 16 373 | 0.001 |
|  | 65-74 | 115 298 | 0.001 | 115 235 | 0.001 |
|  | 75-79 | 1 592 411 | 0.000 | 1 606 341 | 0.000 |
|  | 80-84 | 1 140 399 | 0.000 | 1 150 879 | 0.000 |
|  | 85+ | 1 081 137 | 0.000 | 1 089 493 | 0.000 |
| Prior COVID-19 vaccine count | 2-4 | 526 977 | 0.000 | 529 636 | 0.000 |
|  | 5 | 3 343 190 | 0.000 | 3 373 191 | 0.000 |
|  | 6+ | 75 398 | 0.001 | 75 495 | 0.001 |

#### Table S4b: Testing rates after boosting for the 75+ cohort

| Sub-group |  | Pfizer BA.4-5 | | Sanofi | |
| --- | --- | --- | --- | --- | --- |
|  |  | Person-weeks | All tests | Person-weeks | All tests |
| Main |  | 5 386 441 | 0.000 | 5 434 490 | 0.000 |
| Age | 75-79 | 2 522 642 | 0.000 | 2 545 818 | 0.000 |
|  | 80-84 | 1 560 133 | 0.000 | 1 574 892 | 0.000 |
|  | 85+ | 1 303 666 | 0.000 | 1 313 780 | 0.000 |
| Clinically at-risk | Not clinically at-risk | 1 572 494 | 0.000 | 1 587 776 | 0.000 |
|  | Clinically at-risk | 3 813 947 | 0.000 | 3 846 713 | 0.000 |
| Prior COVID-19 vaccine count | 2-4 | 732 982 | 0.000 | 737 567 | 0.000 |
|  | 5 | 4 584 580 | 0.000 | 4 627 950 | 0.000 |
|  | 6+ | 68 878 | 0.001 | 68 973 | 0.001 |

### Cumulative incidences by subgroup

#### Figure S3a.1: Cumulative incidence estimates per 1,000 people for the CV cohort

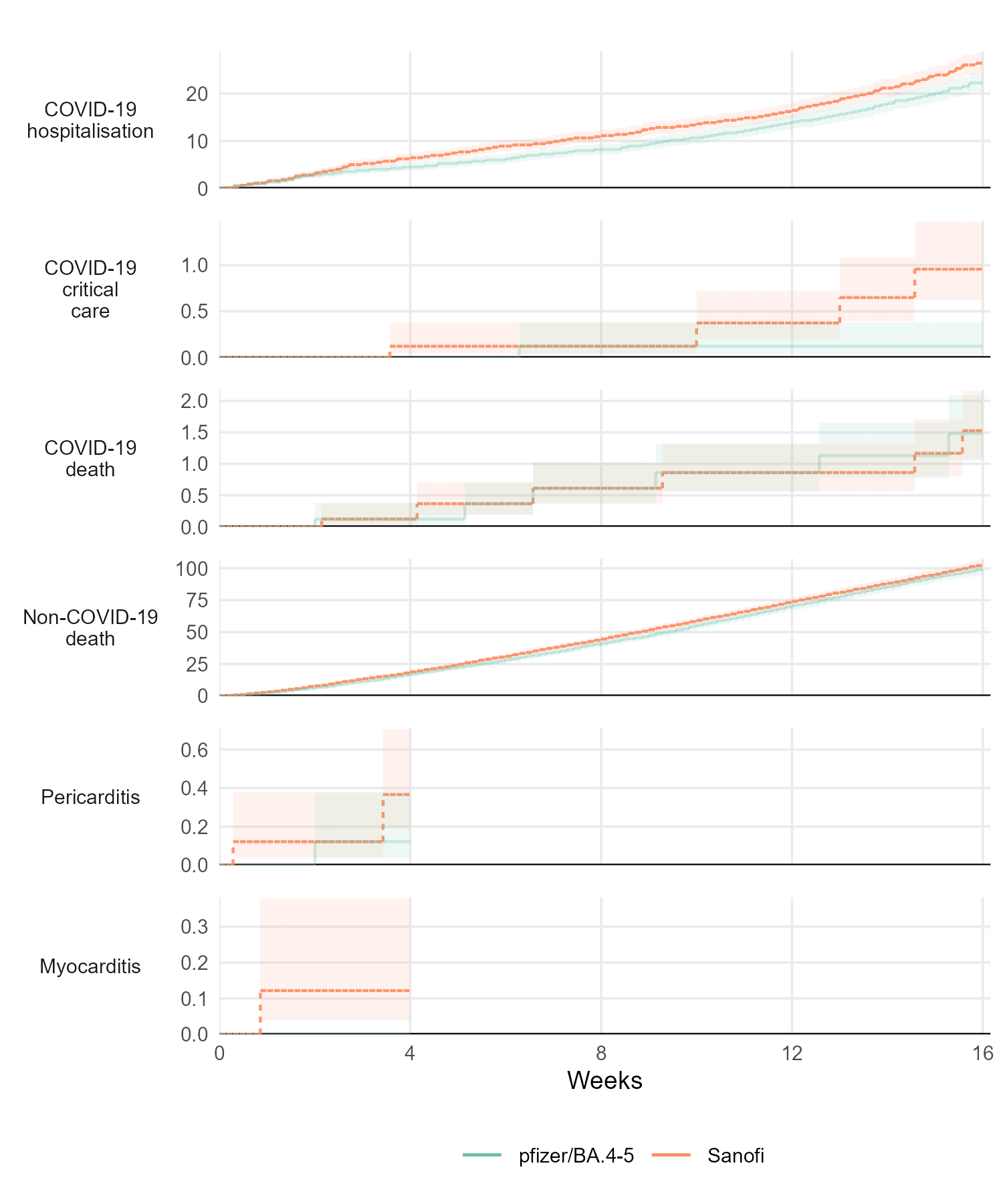

#### Figure S3a.2: Cumulative incidence estimates per 1,000 people by age group for the CV cohort

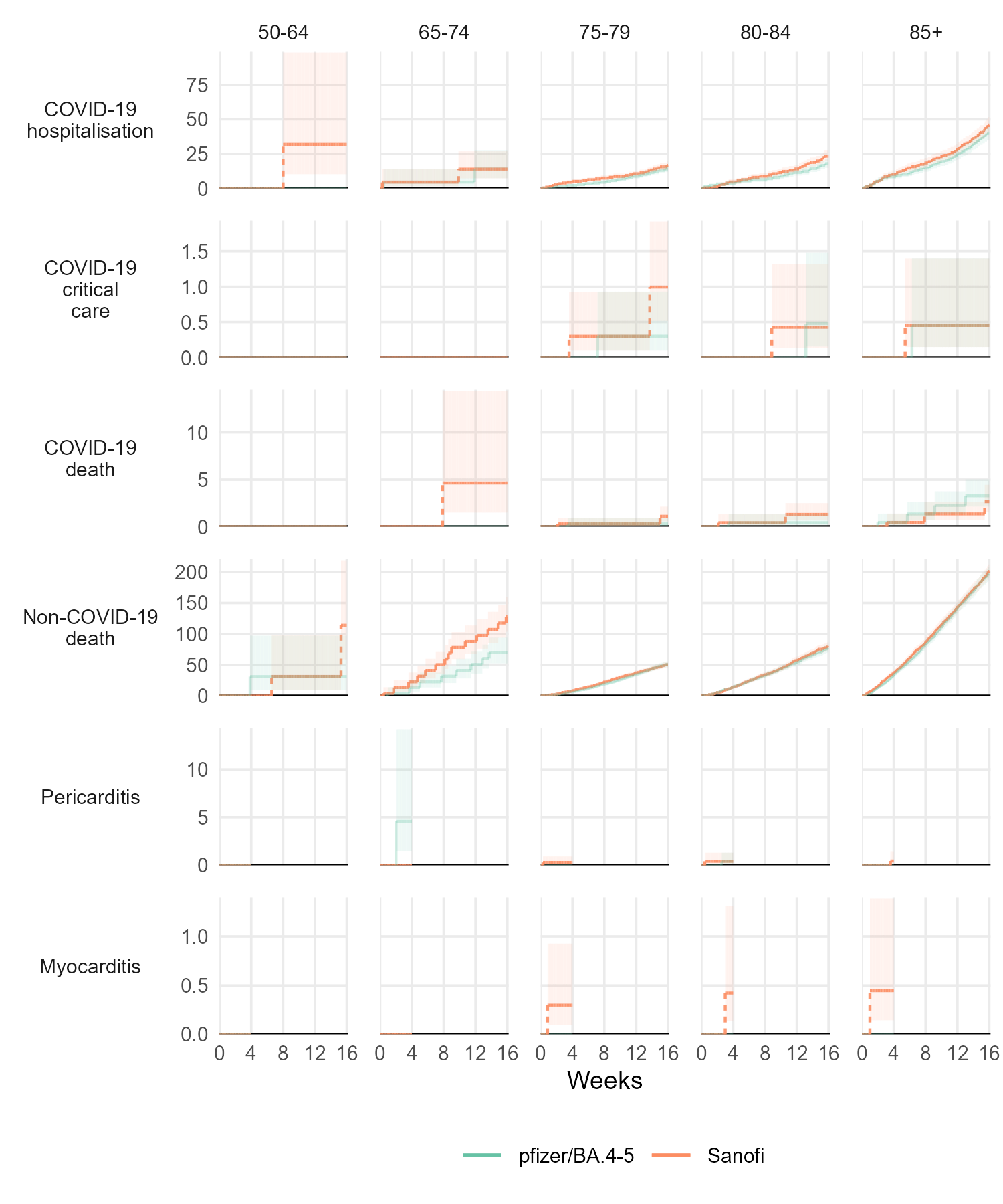

#### Figure S3a.3: Cumulative incidence estimates per 1,000 people by prior vaccine count for the CV cohort

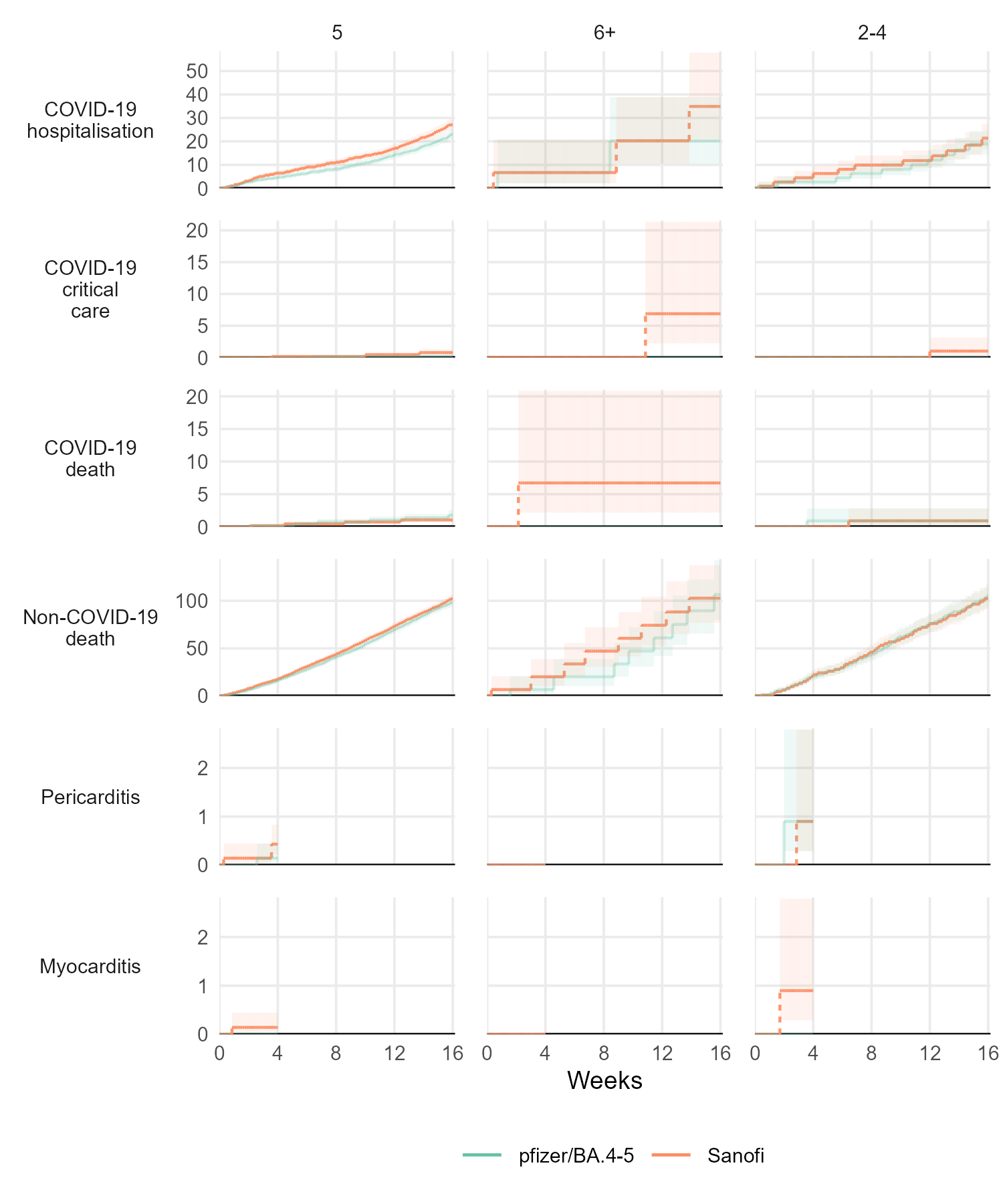

#### Figure S3b.1: Cumulative incidence estimates per 1,000 people for the 75+ cohort

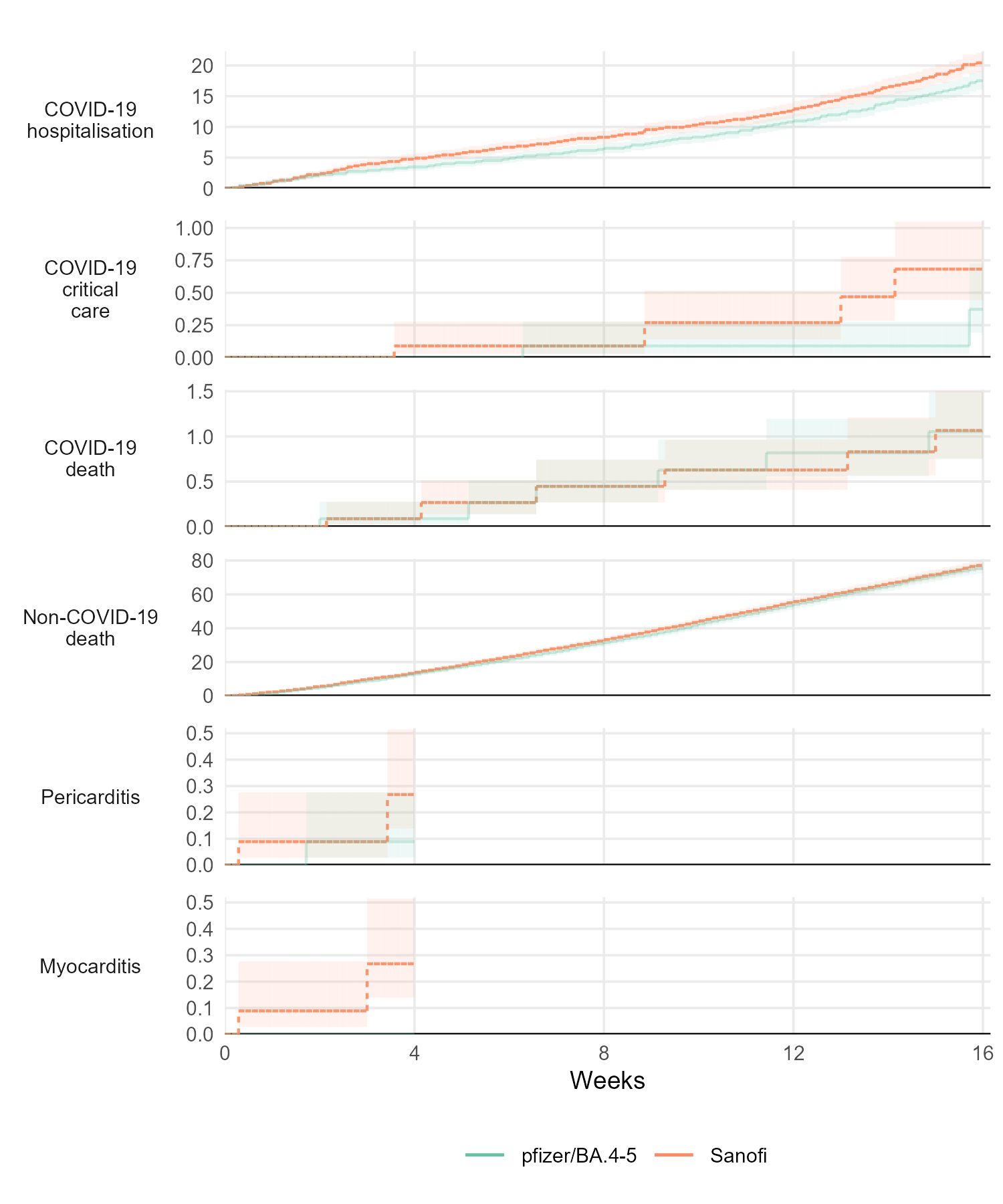

#### Figure S3b.2: Cumulative incidence estimates per 1,000 people by age group for the 75+ cohort

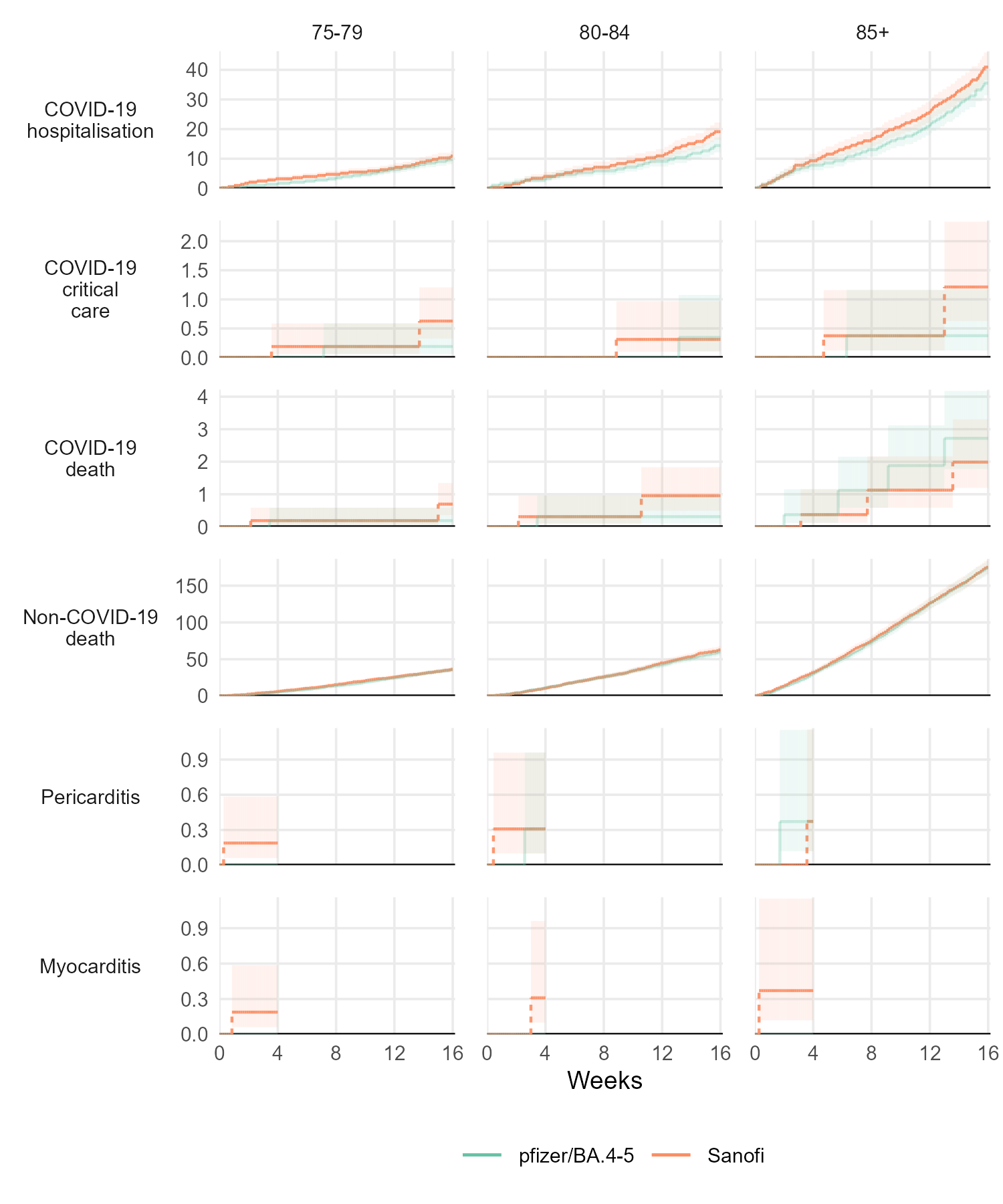

#### Figure S3b.2: Cumulative incidence estimates per 1,000 people by prior vaccine count for the 75+ cohort

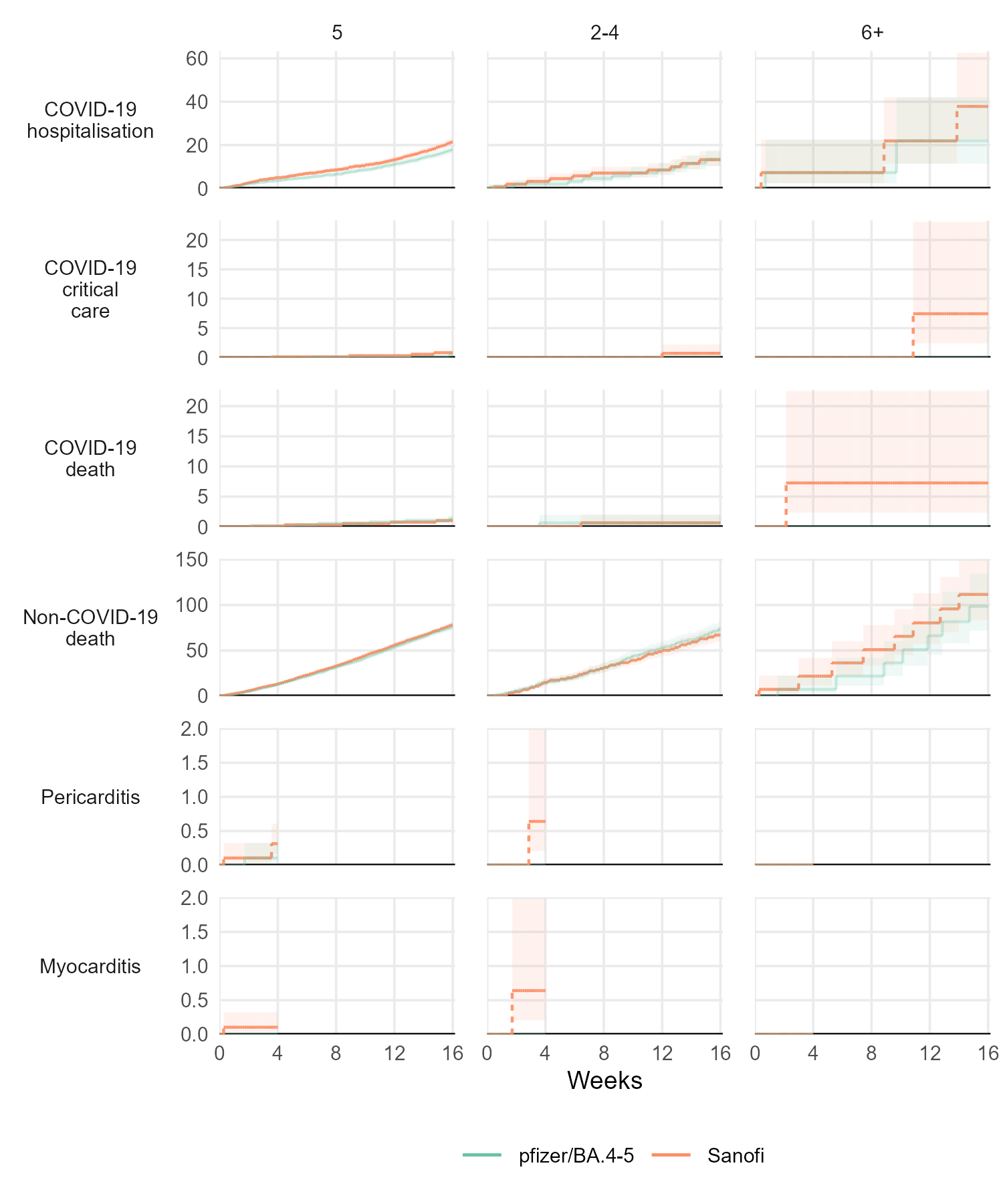

**Figure S3b.4: Cumulative incidence estimates per 1,000 people by clinical vulnerability for the 75+ cohort**
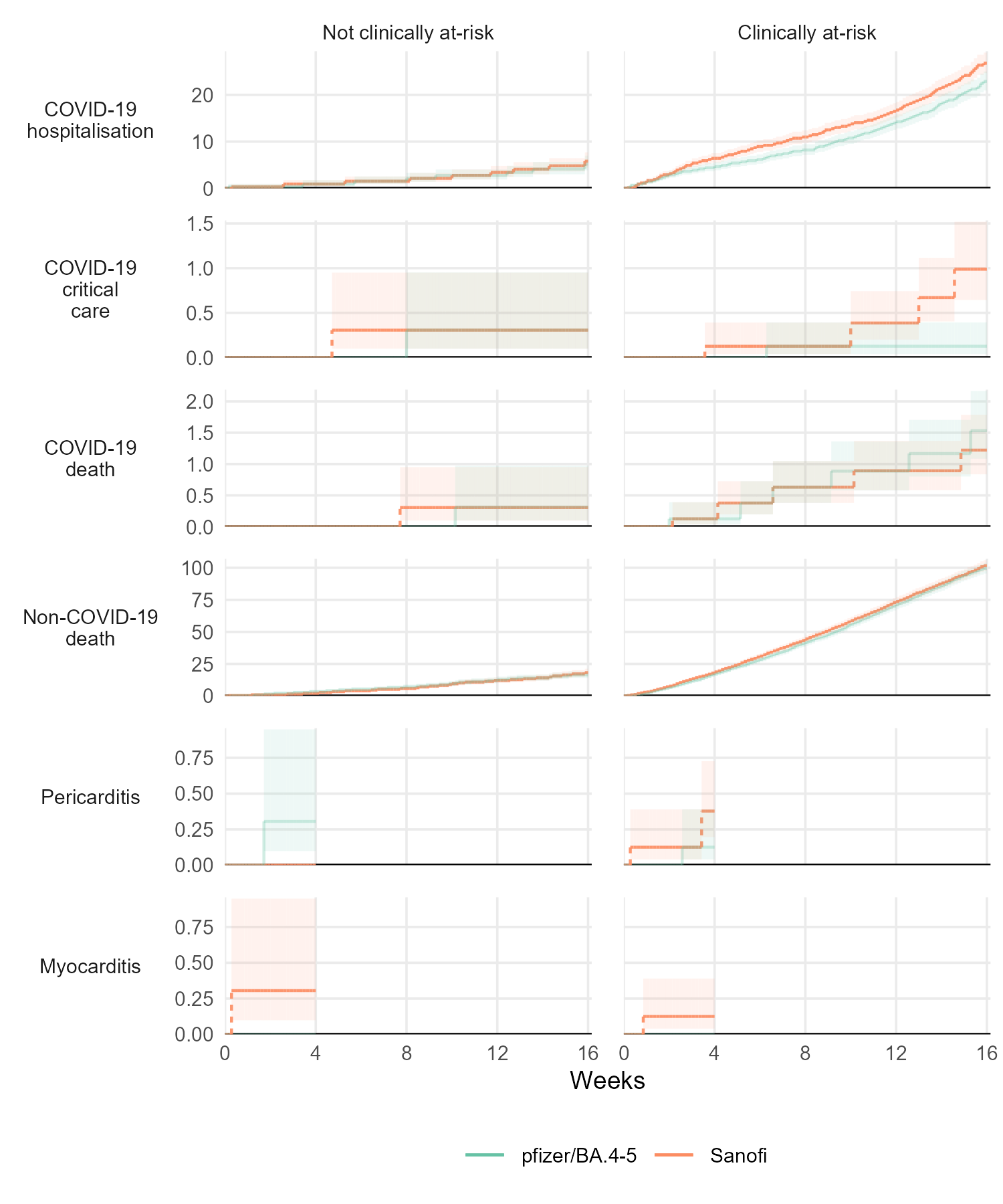

### Comparisons by subgroup

#### Figure S4a.1: Cumulative risk difference, cumulative risk ratio, and period-specific incidence rate ratios for the CV cohort

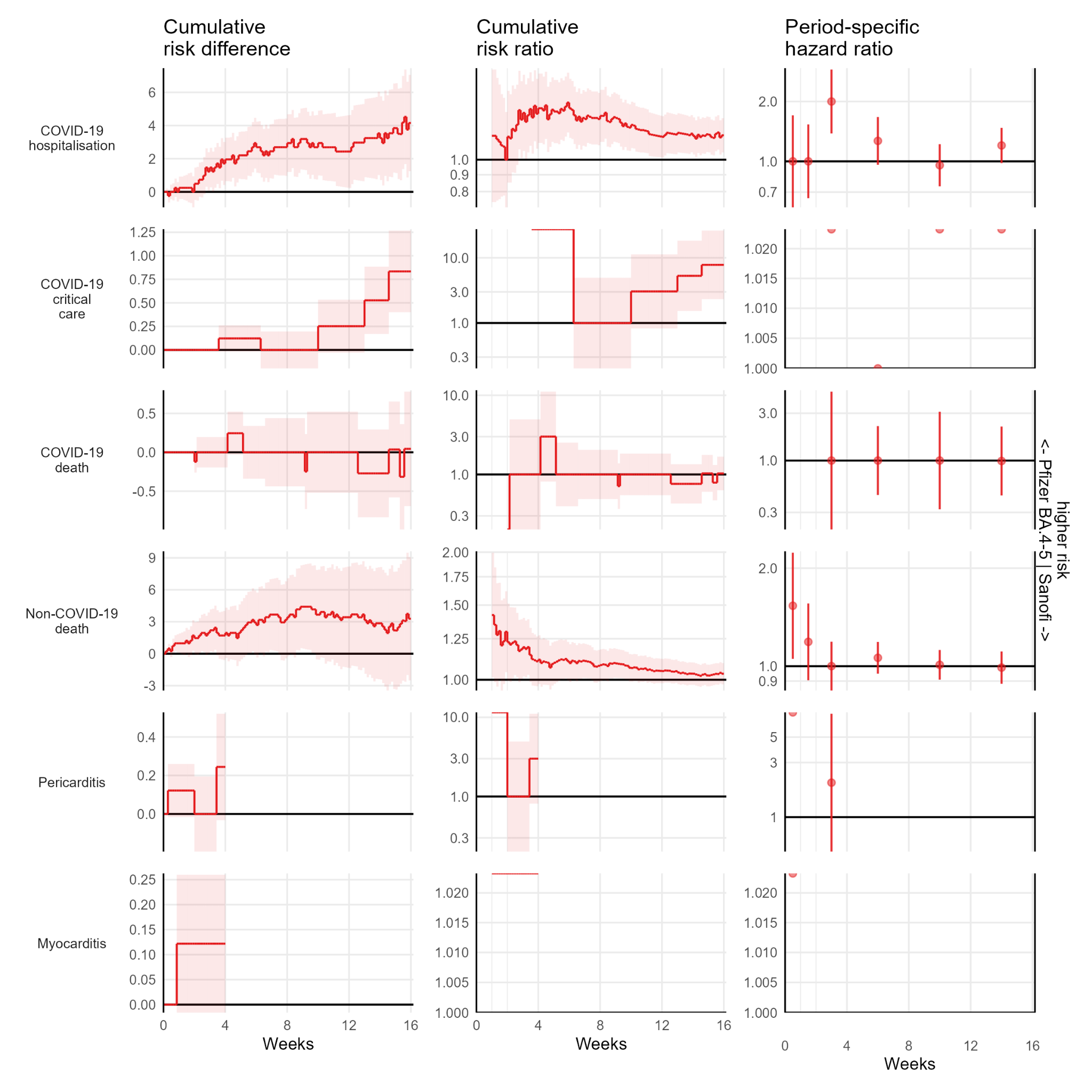

#### Figure S4a.2: Cumulative risk difference, cumulative risk ratio, and period-specific incidence rate ratios by age group for the CV cohort

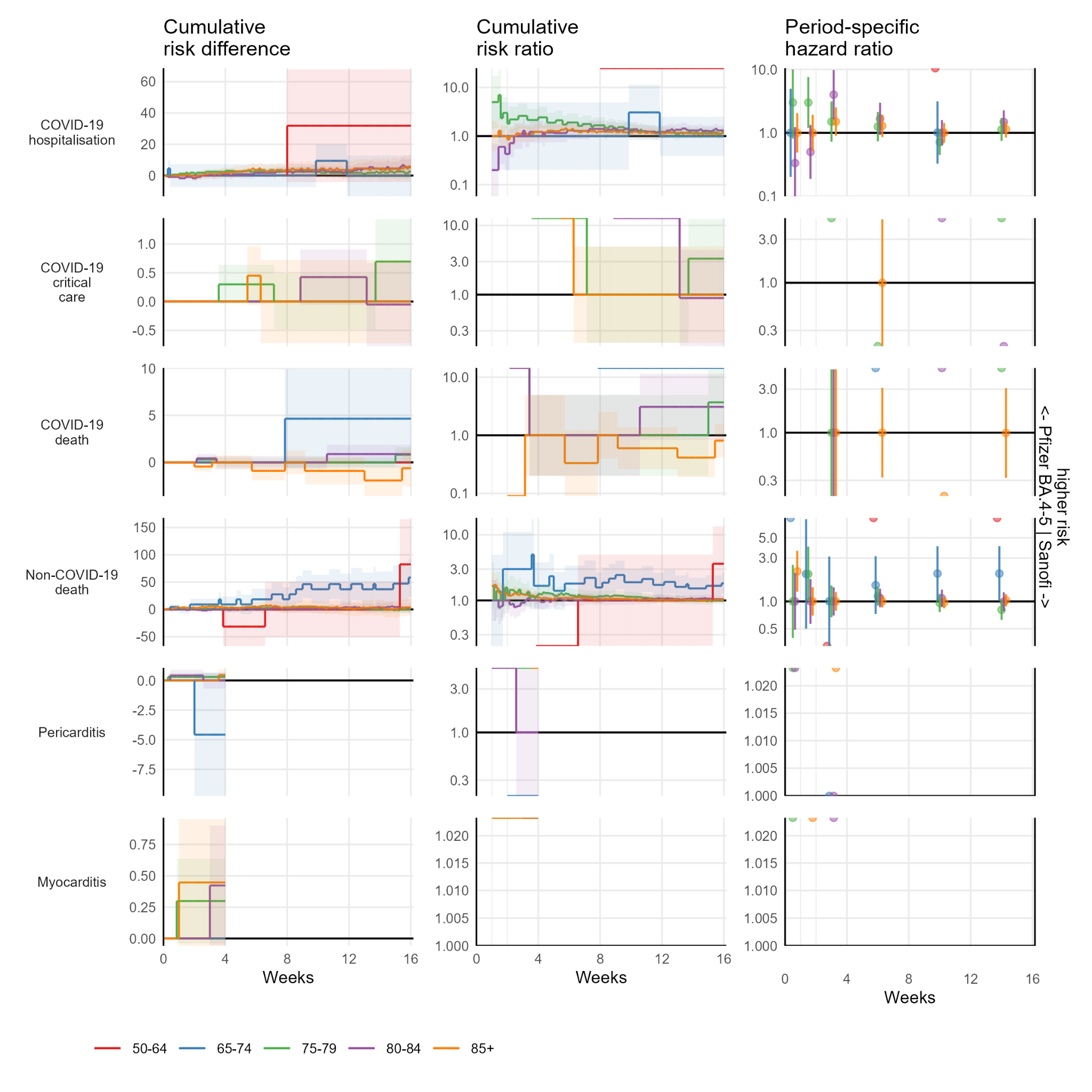

#### Figure S4a.3: Cumulative risk difference, cumulative risk ratio, and period-specific incidence rate ratios by prior vaccine count for the CV cohort

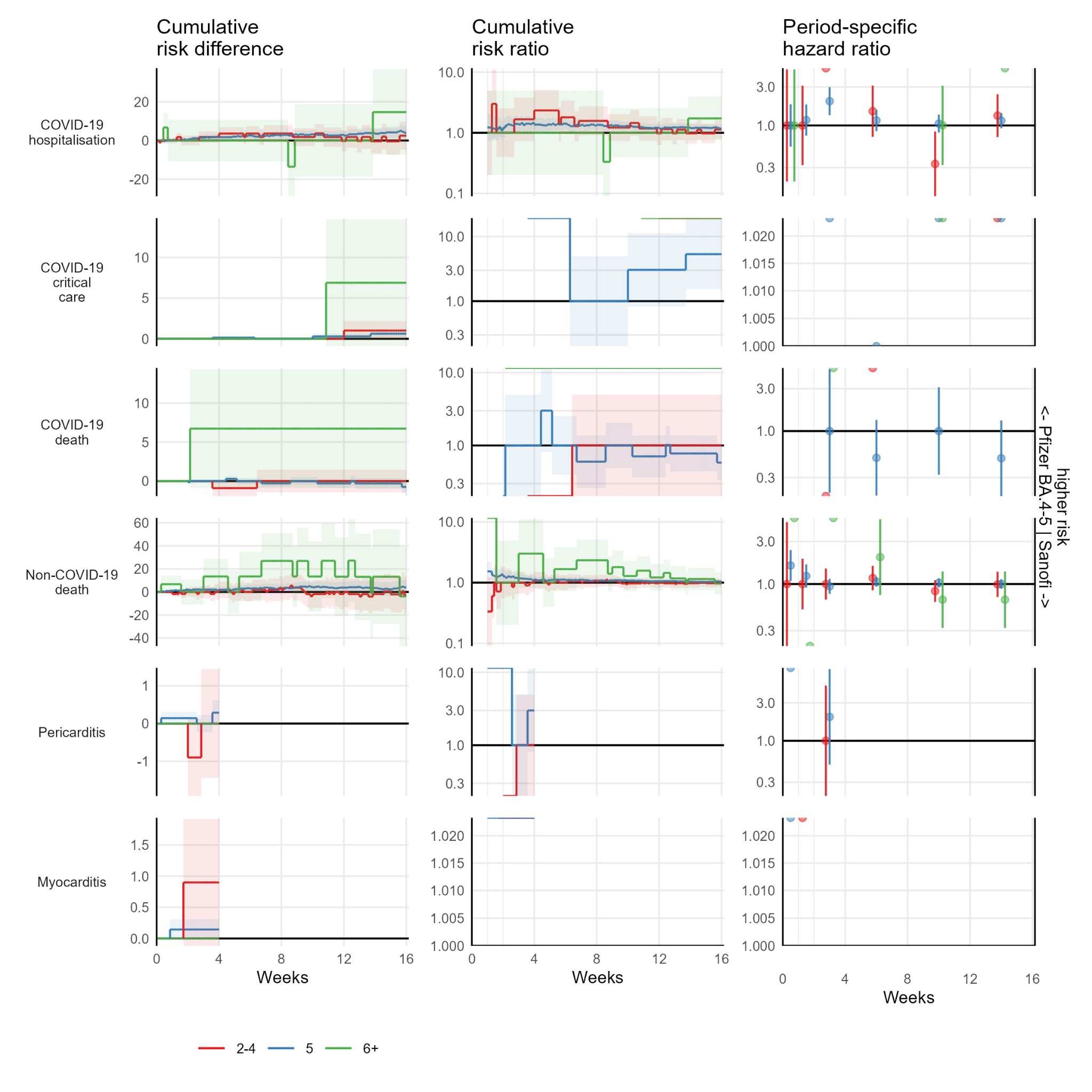

#### Figure S4b.1: Cumulative risk difference, cumulative risk ratio, and period-specific incidence rate ratios for the 75+ cohort

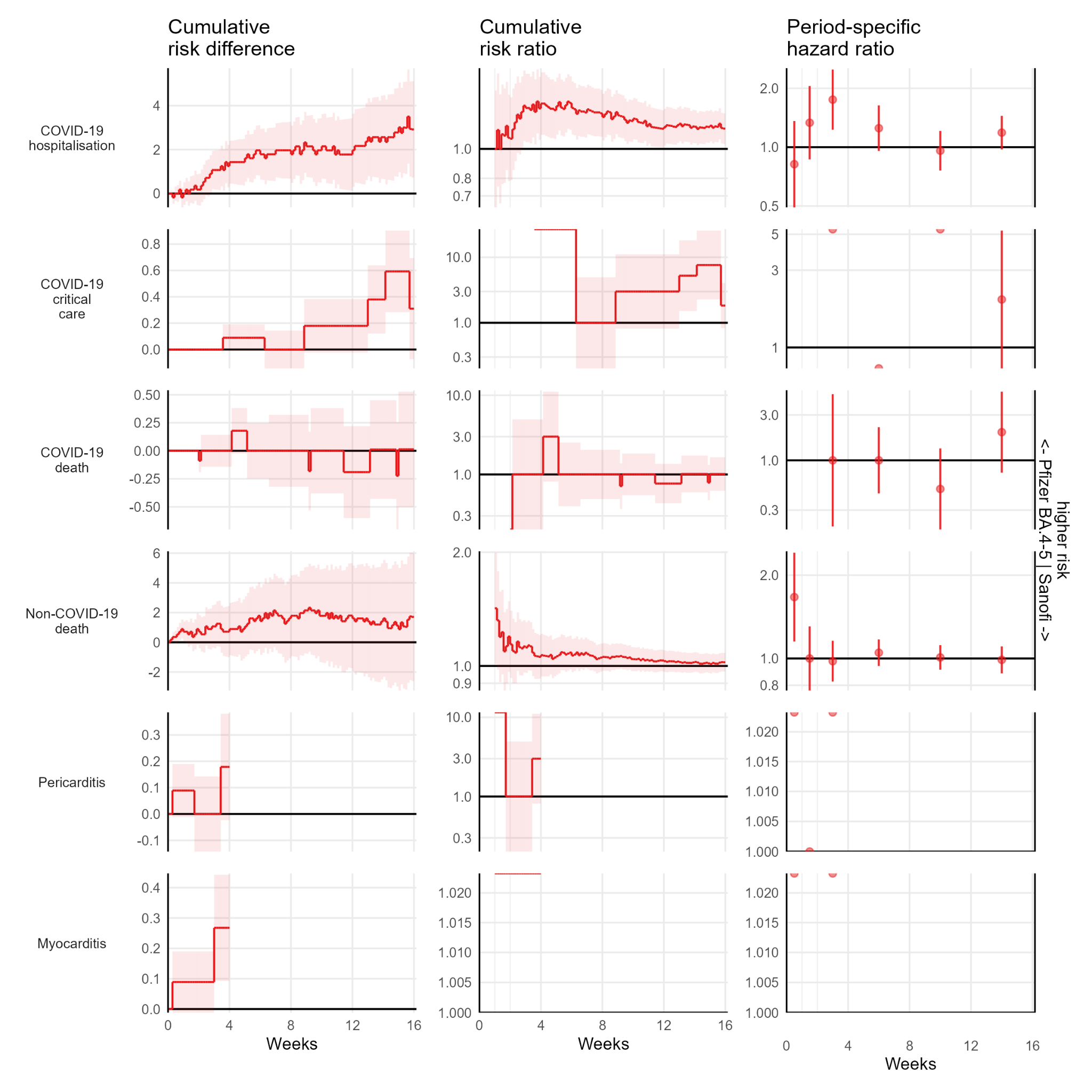

#### Figure S4b.2: Cumulative risk difference, cumulative risk ratio, and period-specific incidence rate ratios by age group for the 75+ cohort

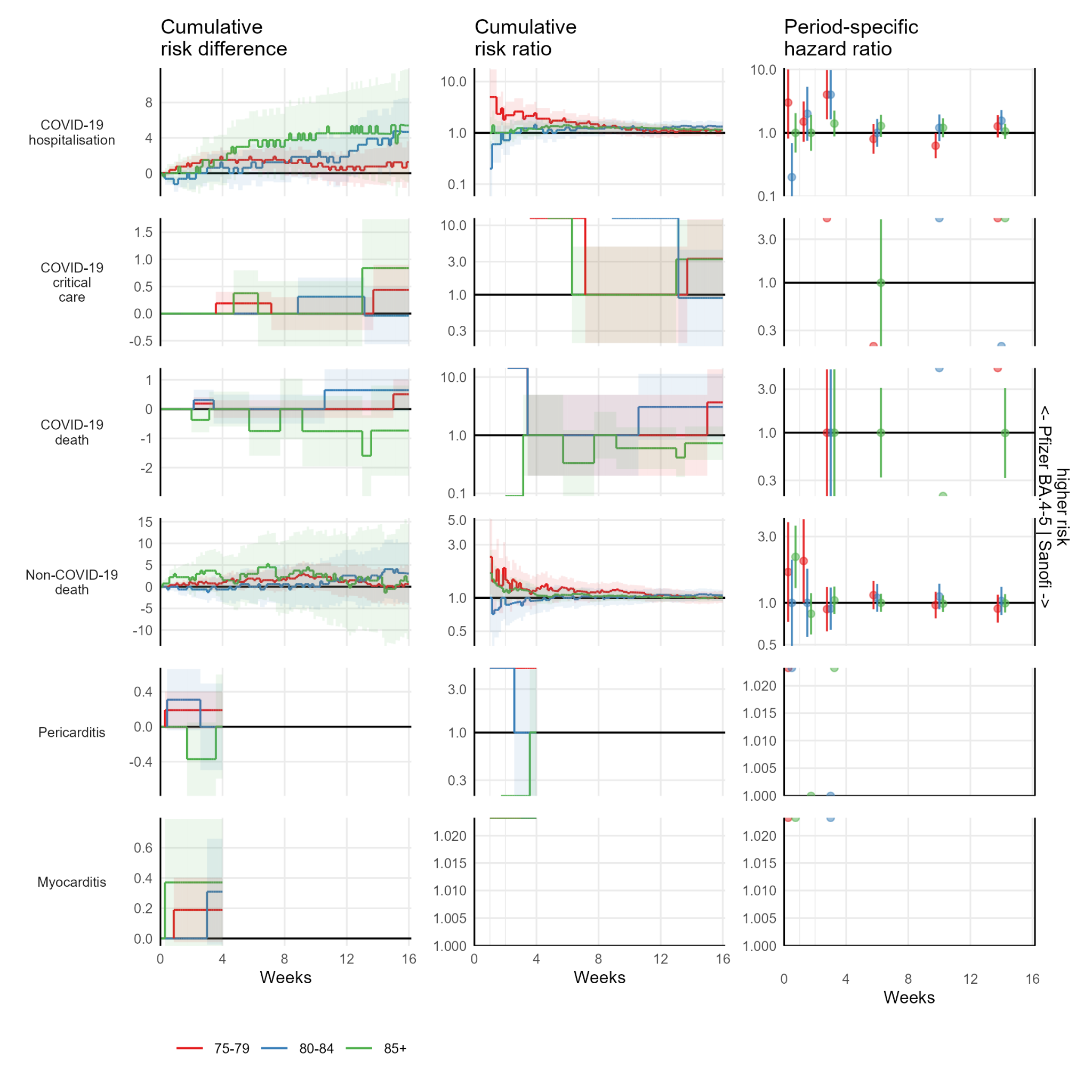

#### Figure S4b.3: Cumulative risk difference, cumulative risk ratio, and period-specific incidence rate ratios by prior vaccine count for the 75+ cohort

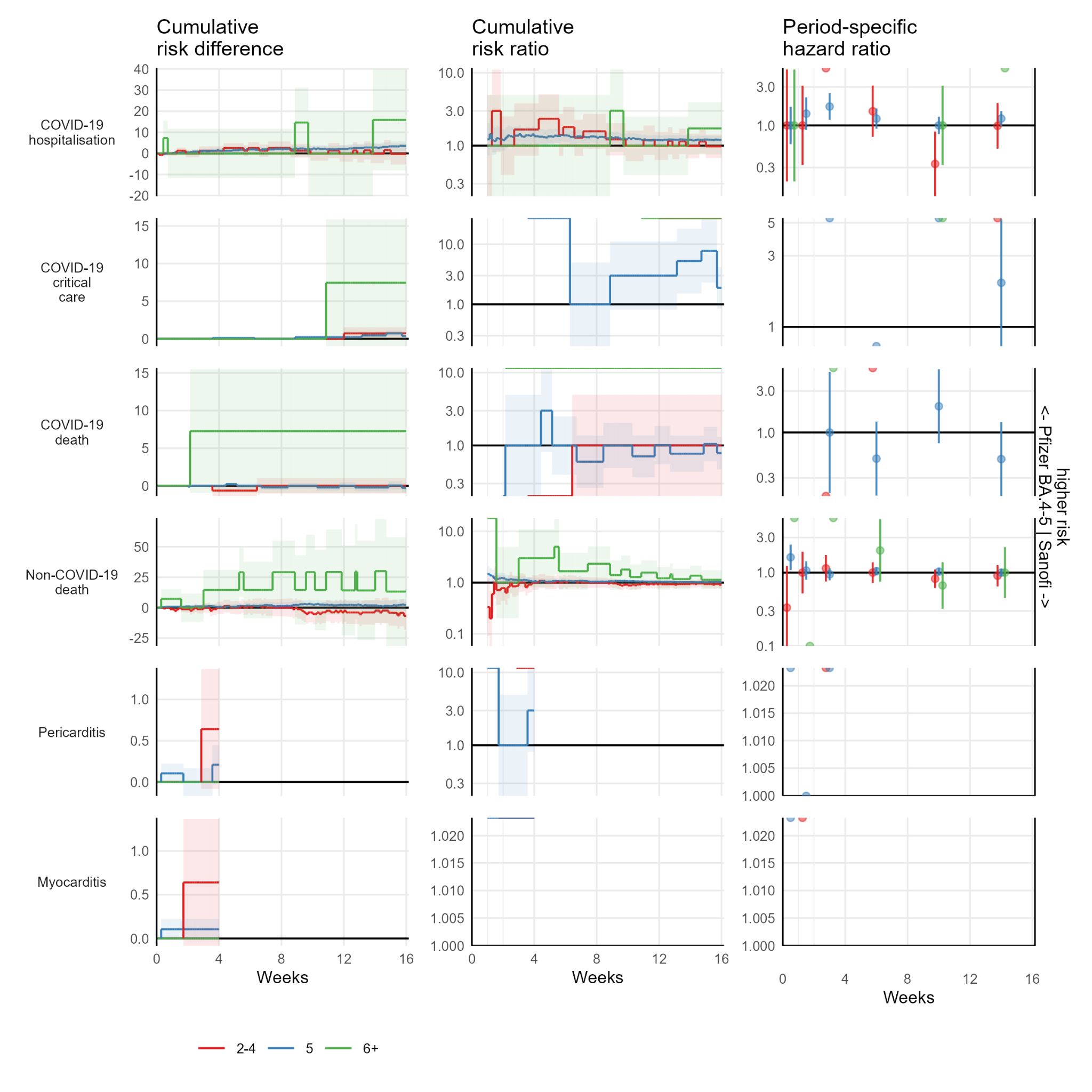

#### Figure S4b.4: Cumulative risk difference, cumulative risk ratio, and period-specific incidence rate ratios by clinical vulnerability for the 75+ cohort

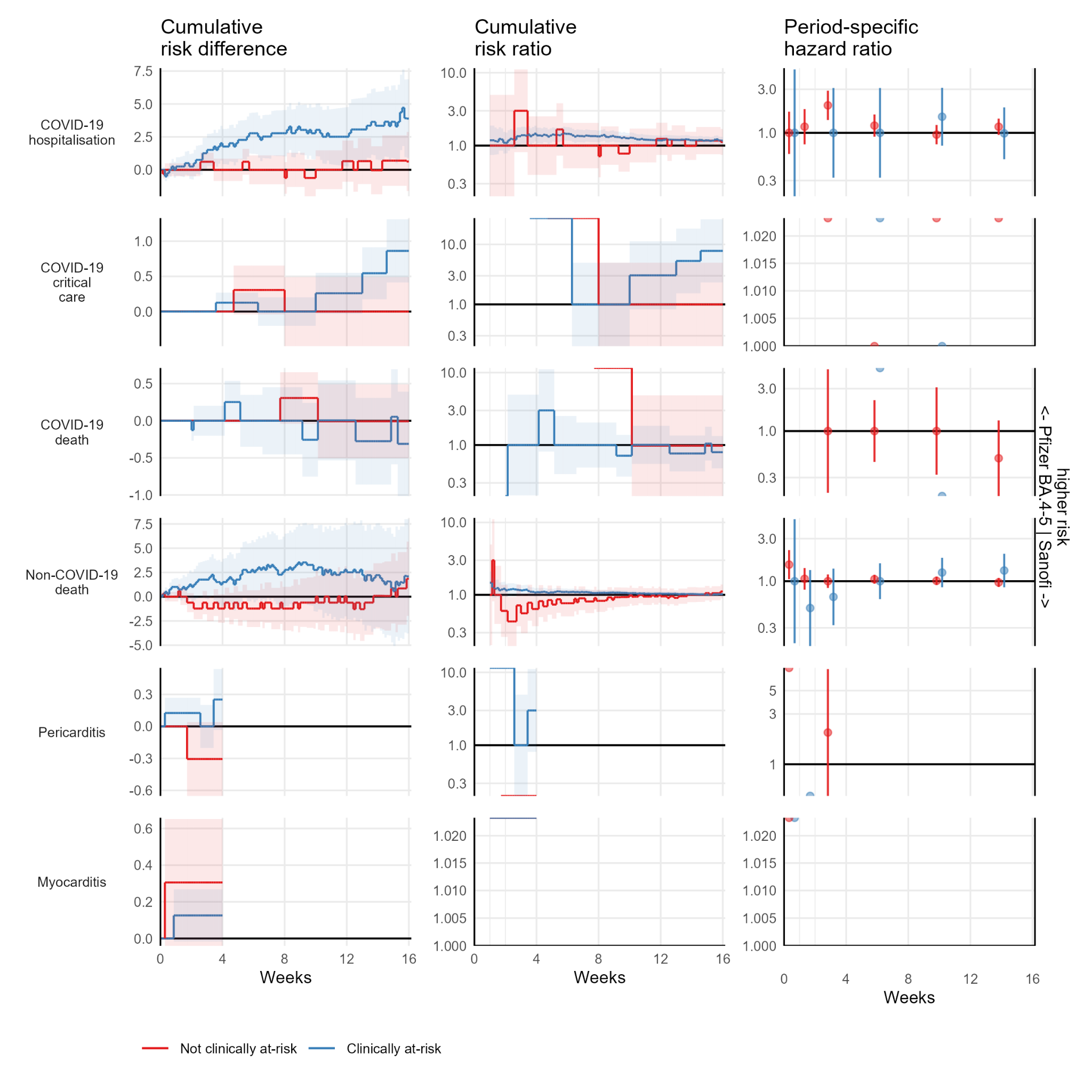
